# Differences in utilization, complications, and mortality after cancer surgery by HIV status among Medicaid beneficiaries from 2001-2021

**DOI:** 10.64898/2026.02.12.26346189

**Authors:** Corinne E. Joshu, Keri Calkins, Jacqueline E. Rudolph, Xiaoqiang Xu, Yiyi Zhou, Maylin Palatino, Karine Yenokyan, Eryka Saylor, Bryan Lau

## Abstract

**Background:** People with HIV (PWH) experience higher cancer-specific mortality and may have worse surgical outcomes than people without HIV (PWoH), though the limited prior evidence largely predates the treat-all antiretroviral therapy (ART) era. We examined postoperative outcomes among PWH and PWoH enrolled in Medicaid in 26 states and Washington, D.C. from 2001–2021.

**Methods:** We identified the first inpatient/outpatient surgery for anal, bladder, breast, colorectal, female genitourinary, gastroesophageal, head and neck, kidney, liver, lung, ovarian, or pancreatic cancer among adults with continuous enrollment for at least 6 months pre- and 3 months post-surgery. Outcomes included length of stay (LOS), 7- and 30-day readmissions (overall and unplanned), emergency department (ED) use, surgical site infection (SSI), and mortality (30-day, 90-day, 1-year, 5-year). Linear, logistic, and Cox proportional hazards models were adjusted for demographics, comorbidities, cancer type, surgical setting and risk, metastasis, and preoperative treatment (radiation/chemotherapy).

**Results:** Among 198,535 beneficiaries undergoing cancer surgery, 4,199 (2.1%) were PWH. PWH were more likely to have inpatient procedures (72.6% vs. 56.4%). Compared to PWoH, PWH had more utilization with longer LOS (7.0 vs. 4.3 days; adjusted mean difference [aMD] = 0.79, 95% CI = 0.60–0.99), extended hospital stays (13.8 vs. 7.4 days; aMD=2.76, 95% CI= 2.42-3.10), and more ED visits (0.82 vs. 0.55 per 90 days; aMD = 0.19, 95% CI = 0.15–0.23). There were no significant differences in readmission, SSI, or 30-day mortality. PWH had higher 90-day mortality (3.2% vs. 1.8%; adjusted odds ratio [aOR] = 1.31, 95% CI = 1.08-1.57), though this was attenuated in the treat-all ART era (2012 – 2021). Results were similar for inpatient surgeries and most common cancer types. PWH had an elevated hazard of 1-year and 5-year mortality post-surgery with an adjusted hazard ratio [aHR] of 1.31 (95% CI = 1.17-1.46) and 1.22 (95% CI= 1.14-1.31), respectively, especially for colorectal cancer (1-year aHR= 1.53, 95% CI=1.24-1.88; 5-year aHR=1.32, 95% CI= 1.14-1.52).

**Conclusions:** PWH had higher post-cancer surgery utilization but similar short-term complications, which supports current guidelines to provide standard cancer care for PWH. More work is needed to elucidate the factors contributing to higher long-term mortality among PWH.

## Background

Cancer is a leading cause of morbidity and mortality among people with HIV (PWH) in the United States in the treat-all era of antiretroviral therapy (ART; post-2012). ^1–4^ PWH can tolerate and should generally receive standard cancer therapy, including surgical treatment, chemotherapy, radiotherapy, and immunotherapy.^5^ Prior studies in the US have reported higher mortality after cancer diagnosis in PWH as compared to PWoH even after accounting for stage at diagnosis, particularly among those with lower CD4 cell counts and AIDS-defining illness,^6–9^ though the exact mechanisms underlying this association is not fully understood. There is also some evidence that PWH may be at risk for elevated rates of short-term and long-term complications as compared to people without HIV (PWoH).

Evidence as to whether PWH have higher risk of short-term complications after cancer surgery is mixed. Prior to the treat-all ART era, PWH undergoing colorectal cancer surgery had increased length of stay (LOS), higher in-hospital mortality during the initial surgery, and higher incidence of wound complications.^10^ An analysis of non-AIDS defining cancers found elevated surgical risk for pulmonary lobectomies and colectomies among PWH.^11^ However, national Veteran’s Affairs data found no significant differences in 30-day or 90-day mortality, wound infection or sepsis, or other post-operative complication after lung cancer surgery from 2000 – 2016, although sample size was limited for PWH.^12^ Data on differences in short-term complications by HIV status for other cancer types/surgeries in the modern ART era remains limited. There is broad evidence that PWH have higher rates of readmission for all hospital stays as compared to PWoH, ^13,14^ though this may be improving slightly over time.^15^ PWH also generally have higher emergency department (ED) utilization overall.^16,17^ However, there is no robust data focused specifically on readmission risk and differences in acute care utilization following cancer surgery for PWH.

We assessed the differences in utilization, surgical complications, and mortality by HIV status following the first observed inpatient or outpatient cancer surgery for 12 common cancer types among Medicaid beneficiaries with and without HIV from 2001 to 2021 in 26 US states and the District of Columbia (DC). These cancer types were included in Centers for Medicare and Medicaid Innovation (CMMI)’s Enhancing Oncology Model, which provides robust claims-based resources to identify cancer treatment, including relevant cancer surgeries.^18^ Outcomes included LOS, total days in hospital within 90 days of surgery, rate of ED visits within 90 days of surgery, rate of surgical site infection (SSI) within 90 days of surgery, 7-and 30-day readmission, 30-day unplanned readmission, and mortality at 30-days, 90-days, 1-year, and 5-years after surgery. Medicaid is the largest payer for HIV care in the US, covering 40% of PWH, yielding substantial longitudinal data on this population, as well as a robust sample size to examine subgroup differences.^19^ We hypothesized that some short-term complications and mortality would be higher for PWH, with results improving in the treat-all ART era (post-2012).^20^

## Methods

### Study Sample

We used enrollment, inpatient, outpatient, and long-term claims data from the Centers for Medicare and Medicaid Services (CMS) for Medicaid beneficiaries aged 18-64 who enrolled between 2001 and 2021 in Washinton, DC or one of 26 states (Alabama, Arkansas, Arizona, California, Colorado, Florida, Georgia, Illinois, Indiana, Kentucky, Maryland, Massachusetts, Michigan, Missouri, Mississippi, Nevada, New Jersey, New York, North Carolina, Ohio, Oklahoma, Pennsylvania, South California, Tennessee, Texas, and Washington). We restricted the study sample to those determined to have one of 12 cancers (anal, bladder, breast, colorectal, female genitourinary, gastroesophageal, head and neck, kidney, liver, lung, ovarian, or pancreatic) based on at least 1 inpatient or 2 outpatient ICD-9-CM or ICD-10-CM diagnosis codes within a 2-year period (Table S1). The Johns Hopkins Bloomberg School of Public Health Institutional Review Board approved this analysis and waived the requirement for informed consent.

Among those with a cancer diagnosis, we then determined whether beneficiaries diagnoses with cancer received surgical treatment. We ascertained the list of ICD-9-PCS, ICD-10-PCS, or CPT codes corresponding to surgical treatment for each of the 12 cancers of interest using the Technical Payment Resources for CMMI’s Enhancing Oncology Model (Table S2).^18^ We identified the first observed surgery corresponding to cancer type on or after the date of cancer diagnosis in inpatient or outpatient claims. To be included in the final analytic sample, beneficiaries surgically treated for cancer were required to be continuously enrolled with full benefits and no dual enrollment in Medicare or private insurance for at least 6 months before surgery to assure complete capture of claims and identify covariates of interest. Beneficiaries were also required to be continuously enrolled for the entirety of the month of surgery and at least 3 months after surgery, unless they died, for a total of 10 months of continuous enrollment. Analytic baseline was the date of the first observed cancer surgery.

### Exposure

Following the CMS Data Warehouse definition for identifying people with HIV, HIV status was identified using at least 1 inpatient or 2 outpatient ICD-9-CM or ICD-10-CM diagnosis codes for HIV within a 2-year period with the first code occurring prior to the first observed cancer surgery (Table S1).^21^

### Covariates

We obtained race/ethnicity (non-Hispanic white, non-Hispanic Black, Hispanic, or other/unknown), sex (male or female), US state, and dates of birth and death from the personal summary file. We defined Charlson Comorbidity Index (CCI) diagnoses^22^ as any ICD-9-CM or ICD-10-CM diagnosis code on an inpatient, outpatient, or long-term care claim in the six months prior to the first cancer surgery date (Table S1). Comorbidity count was categorized as none, 1, or 2 or more comorbidities. Receipt of oral chemotherapy, infusion chemotherapy, or radiation in the six months prior to surgery were identified using HCPCS codes, NDC codes, and a combination of HCPCS, ICD-9-PCS, and ICD-10-PCS codes, respectively, which were also based on the CMMI EOM definitions^18^ (Table S2 and S3).

We used ChatGPT (version 4; OpenAI, 2025) to categorize 1,075 number of possible surgical procedures into high, medium, or low risk using the following prompt: “Categorize the following surgical codes into high, medium or low expected risk based on rates of surgical site infection, in-hospital death, or readmission associated with the procedure”(Table S4). We reviewed the assignment of codes within cancer type to check the face validity of the categorization. For example, resections were generally classified as high risk, while percutaneous procedures were likely to be categorized as low risk.

Metastatic cancer was identified using the presence of any ICD-9-CM or ICD-10-CM diagnosis code for metastatic disease using the Charlson comorbidity index definition^22^ (Table S1) on an inpatient or outpatient claim during the 6 months before cancer surgery.

### Outcomes

Outcomes for the analysis included length of stay (LOS) for the initial cancer surgery and total number of days in hospital within 90 days after surgery, which was a sum of the LOS for the initial surgery and any subsequent readmissions where the admission date was in the 90-day time window. We also identified death within 30 days, 90 days, 1-year, and 5-years after the initial surgery.

We identified admission or readmission after cancer surgery, depending on if the first observed cancer surgery was in the inpatient or outpatient setting, within 7- and 30-days using admission and discharge dates on inpatient claims and dates of service on outpatient claims. We also identified unplanned readmissions within 7 and 30 days using the CMS’s definition used in the Quality Payment Program.^23^

Surgical site infection (SSI) within 90 days after cancer surgery was identified using a previously validated claims-based algorithm.^24^ Finally, we identified the rate of Emergency Department (ED) visits and outpatient observation stays per 1,000 beneficiaries in the 90 days after surgery. We limited ED visits to treat-and-release ED visits and observation stays in the outpatient file and identified using any revenue center codes indicating an ED claim (0450 – 0452, 0456, 0459, and 0981), any CPT code for ED services (99281-99285), any CPT code for a surgical procedure (10004-69979) occurring the ED (place of service code 32), any CPT code for an observation stay (99217-99220, 99224-99226, or 99234-99236), or a HCPCS code for an observation stay (G0378) where the revenue center codes indicated observation (0760 or 0762).^25^

### Statistical Analysis

We used multivariable linear regression models to assess the adjusted mean difference (aMD) in LOS for the first cancer surgery, number of days in hospital in the first 90 days after surgery, and number of ED visits in the first 90 days after surgery (shown as rate per 1,000 persons) and multivariable logistic regression models to assess the adjusted odds ratio (aOR) of overall and unplanned 7 and 30 day readmission, SSI in the first 90 days after surgery, and death in the first 90 days after surgery. Models were adjusted for age, sex, race, state, year of cancer surgery, cancer type, comorbidity count, receipt of additional cancer treatment with separate indicators for oral chemotherapy or immunotherapy, infusion chemotherapy, or radiation, surgical risk, and metastasis.

We used Cox proportional hazards regression to estimate the adjusted hazard ratio (aHR) and 95% CIs of death by HIV status within 1 and 5 years after cancer surgery. Individuals were followed from the date of surgery until death, time of Medicaid disenrollment, whichever came first. Models were adjusted for age, sex, race, state, year of cancer surgery, cancer type, comorbidity count, receipt of additional cancer treatment with separate indicators for oral chemotherapy or immunotherapy, infusion chemotherapy, or radiation, surgical risk, and metastasis. We conducted all analyses overall and stratified by sex, race/ethnicity, cancer type, and surgical setting (i.e., inpatient, outpatient), sample size permitting. All analyses were conducted using R version 4.3.1 (R Foundation, Vienna, Austria, 2023-07-19).

## Results

We identified 198,535 Medicaid beneficiaries aged 18 – 64 who underwent a first observed cancer surgery while enrolled between 2001 and 2021, of which 4,199 had HIV (2.1%) prior to surgery (Table 1). PWH were less likely to be female (63.1% vs. 78.2%) and the majority were Non-Hispanic Black (52.4%), as compared to a plurality of Non-Hispanic White (48.3%) people among those without HIV (PWoH). PWH and PWoH had similar age distributions, reflecting underlying cancer risk with age. Of the 12 cancers included in the analysis, the most common cancer types were breast and colorectal for both PWH and PWoH, though the distribution of the cancers differed by HIV status (Table 1) PWH had similar evidence of metastasis at first observed cancer surgery (12.2% vs. 13.8%), but higher likelihood of inpatient surgery (72.6% vs. 56.4%). Overall, inpatient surgeries were more common than outpatient for all cancers expect breast (Figure S1). Finally, PWH had lower rates of treatment with radiation (16.0% vs. 21.7%), infusion chemotherapy (12.6% vs. 26.1%), and oral chemotherapy or immunotherapy (56.5% vs. 62.6%) than PWoH.

**Table 1.**
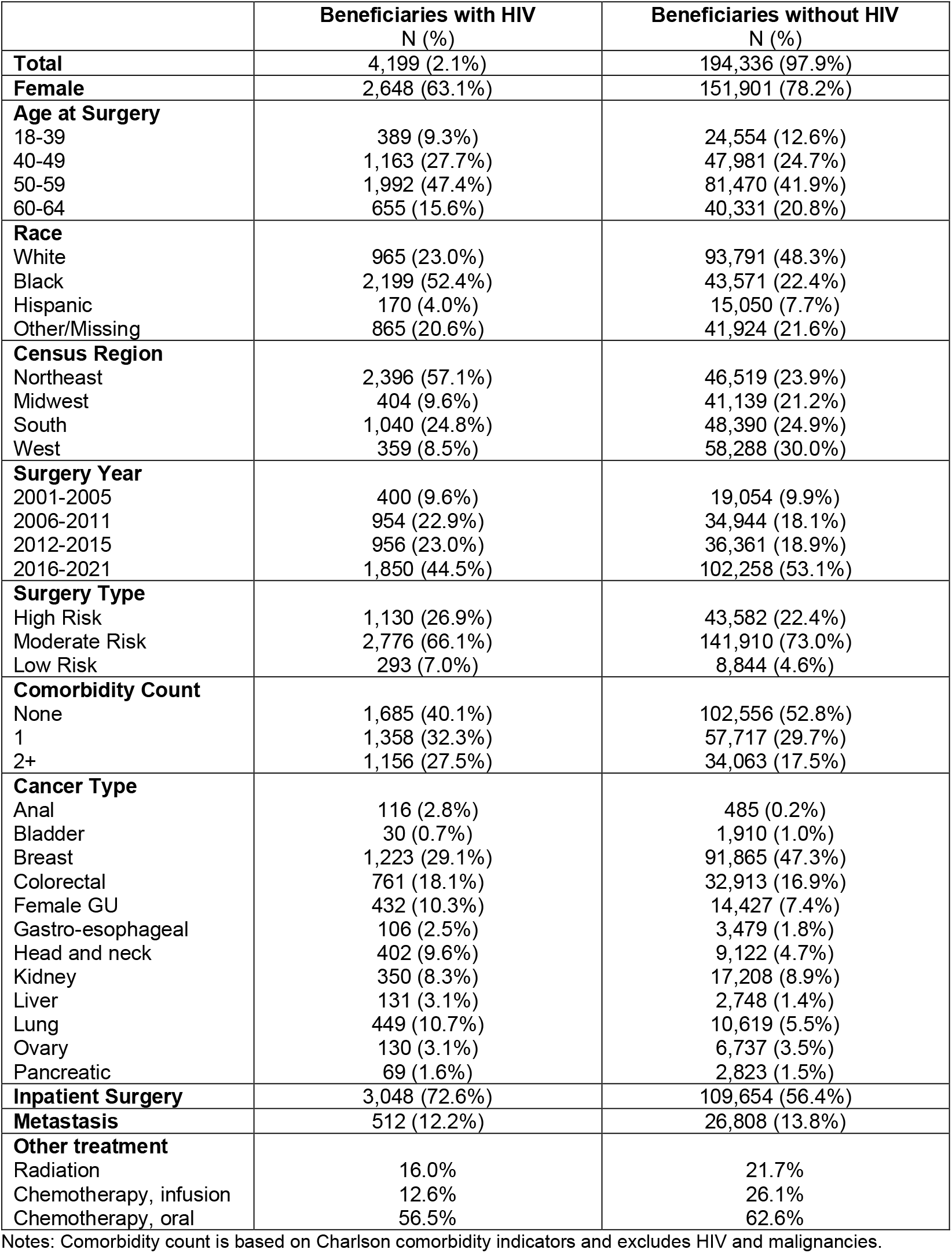
Characteristics of Medicaid beneficiaries with an index cancer-related surgery from 2001-2021 by HIV status.

|  | **Beneficiaries with HIV**<br>N (%) | **Beneficiaries without HIV**<br>N (%) |
| --- | --- | --- |
| **Total** | 4,199 (2.1%) | 194,336 (97.9%) |
| **Female** | 2,648 (63.1%) | 151,901 (78.2%) |
| **Age at Surgery** |  |  |
| 18-39 | 389 (9.3%) | 24,554 (12.6%) |
| 40-49 | 1,163 (27.7%) | 47,981 (24.7%) |
| 50-59 | 1,992 (47.4%) | 81,470 (41.9%) |
| 60-64 | 655 (15.6%) | 40,331 (20.8%) |
| **Race** |  |  |
| White | 965 (23.0%) | 93,791 (48.3%) |
| Black | 2,199 (52.4%) | 43,571 (22.4%) |
| Hispanic | 170 (4.0%) | 15,050 (7.7%) |
| Other/Missing | 865 (20.6%) | 41,924 (21.6%) |
| **Census Region** |  |  |
| Northeast | 2,396 (57.1%) | 46,519 (23.9%) |
| Midwest | 404 (9.6%) | 41,139 (21.2%) |
| South | 1,040 (24.8%) | 48,390 (24.9%) |
| West | 359 (8.5%) | 58,288 (30.0%) |
| **Surgery Year** |  |  |
| 2001-2005 | 400 (9.6%) | 19,054 (9.9%) |
| 2006-2011 | 954 (22.9%) | 34,944 (18.1%) |
| 2012-2015 | 956 (23.0%) | 36,361 (18.9%) |
| 2016-2021 | 1,850 (44.5%) | 102,258 (53.1%) |
| **Surgery Type** |  |  |
| High Risk | 1,130 (26.9%) | 43,582 (22.4%) |
| Moderate Risk | 2,776 (66.1%) | 141,910 (73.0%) |
| Low Risk | 293 (7.0%) | 8,844 (4.6%) |
| **Comorbidity Count** |  |  |
| None | 1,685 (40.1%) | 102,556 (52.8%) |
| 1 | 1,358 (32.3%) | 57,717 (29.7%) |
| 2+ | 1,156 (27.5%) | 34,063 (17.5%) |
| **Cancer Type** |  |  |
| Anal | 116 (2.8%) | 485 (0.2%) |
| Bladder | 30 (0.7%) | 1,910 (1.0%) |
| Breast | 1,223 (29.1%) | 91,865 (47.3%) |
| Colorectal | 761 (18.1%) | 32,913 (16.9%) |
| Female GU | 432 (10.3%) | 14,427 (7.4%) |
| Gastro-esophageal | 106 (2.5%) | 3,479 (1.8%) |
| Head and neck | 402 (9.6%) | 9,122 (4.7%) |
| Kidney | 350 (8.3%) | 17,208 (8.9%) |
| Liver | 131 (3.1%) | 2,748 (1.4%) |
| Lung | 449 (10.7%) | 10,619 (5.5%) |
| Ovary | 130 (3.1%) | 6,737 (3.5%) |
| Pancreatic | 69 (1.6%) | 2,823 (1.5%) |
| **Inpatient Surgery** | 3,048 (72.6%) | 109,654 (56.4%) |
| **Metastasis** | 512 (12.2%) | 26,808 (13.8%) |
| **Other treatment** |  |  |
| Radiation | 16.0% | 21.7% |
| Chemotherapy, infusion | 12.6% | 26.1% |
| Chemotherapy, oral | 56.5% | 62.6% |

Notes: Comorbidity count is based on Charlson comorbidity indicators and excludes HIV and malignancies.

### LOS and days in hospital

Across 2001 – 2021 among all cancer types and surgical settings, PWH had an increased LOS for the initial cancer surgery (7.01 vs. 4.31 days) and increased total days in hospital within the first 90 days after surgery (13.77 vs. 7.44 days) as compared to PWoH (Table 2). Results were similar when limiting to the treat-all ART era (2012 – 2021); PWH spent over 17 days in hospital within the first 90 days after inpatient surgeries as compared to 11 days for PWoH (Table 2). In site-specific analyses, PWH surgically treated for colorectal, head and neck, and lung cancers had significantly longer length of stays and total days in the hospital than PWoH; there was also a modest, but significant difference for PWH surgically treated for breast cancer (Figure 1). Total days in hospital were significantly higher for PWH as compared to PWoH for all six cancer types individually assessed with the largest differences observed for colorectal, head and neck, and lung cancers.

**Table 2.** Complication rates after first observed cancer surgery among Medicaid beneficiaries from 2001-2021 by HIV status, year, and surgical setting.

| Surgery Type | Complication | Frequency |  | Mean Difference or Odds Ratio (95% CI) |
| --- | --- | --- | --- | --- |
|  |  | HIV | No HIV |  |
| Overall |  | N=4,199 | N=194,336 |  |
|  | Length of stay (Mean [SD]) | 7.01 | 4.31 | **0.79 days (0.60, 0.99)** |
|  | Total days in hospital (Mean [SD]) | 13.77 | 7.44 | **2.76 days (2.42, 3.10)** |
|  | 7-day all-cause readmission (%) | 8.9% | 5.8% | 0.97 (0.86, 1.08) |
|  | 30-day all-cause readmission (%) | 18.4% | 12.8% | 1.05 (0.96, 1.14) |
|  | Surgical Site Infection (%)* | 6.1% | 4.2% | 1.08 (0.94, 1.23) |
|  | 30-day mortality (%) | 0.91% | 0.68% | 0.99 (0.71, 1.38) |
|  | 90-day mortality (%) | 3.2% | 1.8% | **1.31 (1.08, 1.57)** |
|  | 30-day unplanned readmission (%)* | 17.3% | 12.0% | 1.03 (0.95, 1.12) |
|  | ED stays (Mean [SD]) | 0.823 | 0.552 | **0.193 stays (0.154, 0.232)** |
| Treat-all ART Era<br>(2012 – 2021) |  | N=2,806 | N=138,619 |  |
|  | Length of stay (Mean [SD]) | 6.68 | 3.95 | **0.82 days (0.59, 1.04)** |
|  | Total days in hospital (Mean [SD]) | 13.28 | 6.70 | **2.98 days (2.59, 3.38)** |
|  | 7-day all-cause readmission (%) | 8.3% | 4.8% | **1.27 (1.09, 1.47)** |
|  | 30-day all-cause readmission (%) | 17.2% | 11.0% | **1.23 (1.11, 1.37)** |
|  | Surgical Site Infection (%)* | 4.6% | 3.1% | 1.05 (0.87, 1.27) |
|  | 30-day mortality (%) | 0.82% | 0.64% | 0.85 (0.55, 1.31) |
|  | 90-day mortality (%) | 3.1% | 1.8% | 1.22 (0.97, 1.54) |
|  | 30-day unplanned readmission (%)* | 15.6% | 10.0% | **1.21 (1.09, 1.36)** |
|  | ED stays (Mean [SD]) | 0.899 | 0.575 | **0.237 stays (0.188, 0.287)** |
| Inpatient Surgeries Only |  | N=3,048 | N=109,654 |  |
|  | Length of stay (Mean [SD]) | 9.28 | 6.87 | **1.04 days (0.74, 1.33)** |
|  | Total days in hospital (Mean [SD]) | 17.28 | 11.43 | **3.06 days (2.56, 3.56)** |
|  | 7-day all-cause readmission (%) | 10.7% | 7.9% | 0.96 (0.85, 1.09) |
|  | 30-day all-cause readmission (%) | 21.5% | 16.8% | 1.03 (0.94, 1.13) |
|  | Surgical Site Infection (%) | 7.4% | 6.2% | 1.06 (0.92, 1.22) |
|  | 30-day mortality (%) | 3.0% | 2.7% | 1.02 (0.73, 1.43) |
|  | 90-day mortality (%) | 4.1% | 2.9% | **1.30 (1.07, 1.58)** |
|  | 30-day unplanned readmission (%) | 20.3% | 15.9% | 1.02 (0.92, 1.12) |
|  | ED stays (Mean [SD]) | 0.94 | 0.70 | **0.216 stays (0.162, 0.269)** |
| Outpatient Surgeries Only |  | N=1,151 | N=84,682 |  |
|  | Length of stay (Mean [SD]) | N/A | N/A | - |
|  | Total days in hospital (Mean [SD]) | 4.47 | 2.27 | **1.61 days (1.33, 1.88)** |
|  | 7-day all-cause readmission (%) | 4.2% | 3.0% | 0.98 (0.72, 1.33) |
|  | 30-day all-cause readmission (%) | 10.2% | 7.7% | 1.09 (0.89, 1.33) |
|  | Surgical Site Infection (%) | 2.4% | 1.6% | 1.42 (0.96, 2.09) |
|  | 30-day mortality (%) | 0.09% | 0.12% | 0.47 (0.06, 3.46) |
|  | 90-day mortality (%) | 0.8% | 0.4% | 1.29 (0.64, 2.60) |
|  | 30-day unplanned readmission (%) | 9.3% | 7.0% | 1.07 (0.87, 1.32) |
|  | ED stays (Mean [SD]) | 0.51 | 0.35 | **0.133 stays (0.077, 0.189)** |

Notes: Adjusts for age, sex, race, state, year of cancer surgery, cancer type, comorbidity count, receipt of additional cancer treatment with separate indicators for oral chemotherapy or immunotherapy, infusion chemotherapy, or radiation, surgical risk, and metastasis

**Figure 1.**
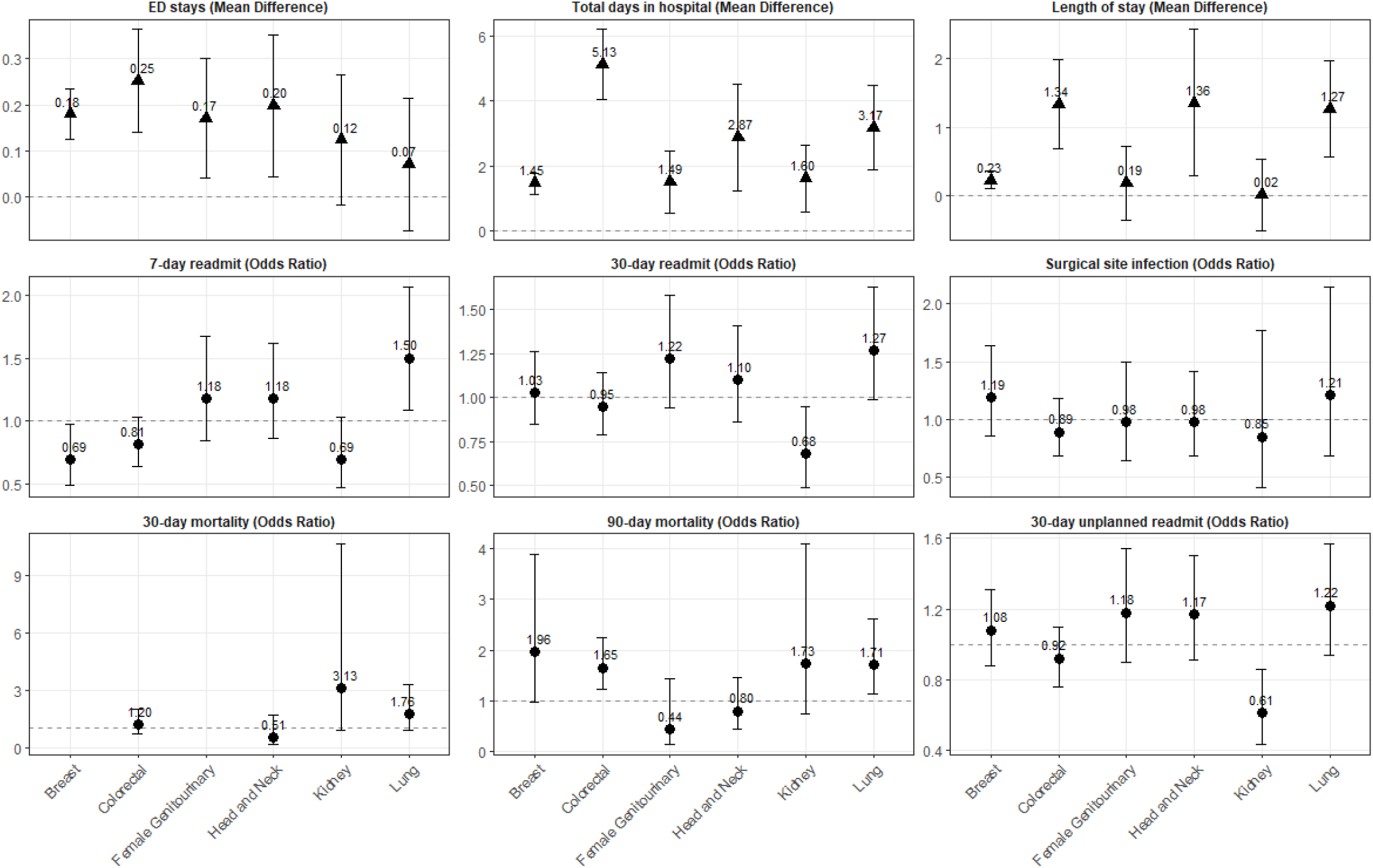
Complication rates after an index cancer-related surgery among Medicaid beneficiaries from 2001-2021 by HIV status and cancer type. Notes: Adjusts for age, sex, race, state, year of cancer surgery, cancer type, comorbidity count, receipt of additional cancer treatment with separate indicators for oral chemotherapy or immunotherapy, infusion chemotherapy, or radiation, surgical risk, and metastasis.

### ED visits

Overall, PWH had a higher number of ED visits within the 90 days after first observed cancer surgery, with a mean of 0.823 visits among PWH versus 0.522 visits among PWoH (aMD=0.193 stays [95% CI=0.154, 0.232]) (Table 2). Results were consistent when limiting to 2012 – 2021, among inpatient surgeries, among outpatient surgeries, and stratified by cancer types, although kidney and lung cancer differences were not statistically significant (Figure 1).

### Readmission

Despite increased number of days in hospital, PWH did not have higher odds of all-cause or unplanned readmission across all surgeries. Interestingly, when limited to the treat-all ART era, PWH were at increased odds of 7-day all-cause readmission, 30-day all-cause readmission, and 30-day unplanned readmission. This difference appears to be at least partly driven by slightly lower readmission rates among PWoH in Treat-all Era compared with the overall time period, relative to PWH. For example, unplanned 30-day readmission among PWH was 17.3% in 2001–2021 and 15.6% in the treat-all ART era, reflecting a 1.7 percentage point (p.p.) improvement, as compared to a 2.0 p.p. improvement of 2.0 p.p. for PWoH (12.0% to 10.0%), resulting in PWH having 1.2 times the odds of 30 day readmission compared to PWoH in the 2012-2021 time period(95% CI=1.09, 1.36, Table 2). Results were not significantly different when limiting to inpatient surgeries. In site specific analyses, 7-day readmission was lower for PWH surgically treated for breast cancer as compared to PWoH, but there was no difference in 30-day readmission (Figure 1). PWH surgically treated for kidney cancer had non-statistically significant lower odds of 7-day readmission and significantly lower odds of 30-day readmission as compared to their PWoH counterparts. In contrast, PWH surgically treated for lung cancer had higher odds of 7-day readmission and non-statistically significant higher odds of 30-day readmission as compared to their PWoH counterparts. There were no differences in readmission by HIV status for colorectal, female genitourinary, or head neck cancers.

### Surgical Site Infection

PWH had similar odds of SSI by 90 days as PWoH overall (6.1% vs. 4.2) and among both inpatient (7.4% vs. 6.2) and outpatient surgeries (2.4% vs. 1.6%; Table 2). SSI was lower for both groups in the treat-all ART era with no significant differences observed by HIV status (4.6% vs. 3.1%). No significant differences were observed among the six cancer types examined (Figure 1).

### Short-term mortality

There were no significant differences in 30-day mortality overall, in the treat-all ART era, or among inpatient or outpatient surgeries. In site-specific cancer analyses, PWH surgically treated for kidney or lung cancer appeared to have higher odds of 30-day mortality as compared to their PWoH counterparts, however, these were not statistically significant (Figure 1). There were too few deaths within 30-days of breast cancer surgery to compare odds by HIV status. There was evidence of higher 90-day mortality among PWH overall, which was appeared to be driven by inpatient surgeries (Table 2). Differences were no longer statistically significant in the treat-all ART era. In site-specific analyses, 90-day mortality was significantly higher for PWH undergoing surgery for colorectal and lung cancer, and appear slightly elevated for breast and kidney cancer, but differences were not statistically significant.

### Long-term mortality

Overall, PWH had poorer 5-year crude survival than PWoH after surgical treatment for cancer (Figure 2). After adjustment, PWH had significantly higher mortality at 1- and 5-years after surgery than PWoH, overall and in the treat-all ART era (Figure 3). These differences were significant for inpatient surgeries but not outpatient surgeries. In site specific analyses, mortality was significantly higher for PWH at 1-year after surgical treatment for colorectal cancer at compared to those without HIV. As compared to PWoH, PWH had significantly higher mortality 5 years lung, female genitourinary, and kidney cancers but not breast or head-and-neck cancers (Figure 3).

**Figure 2.**
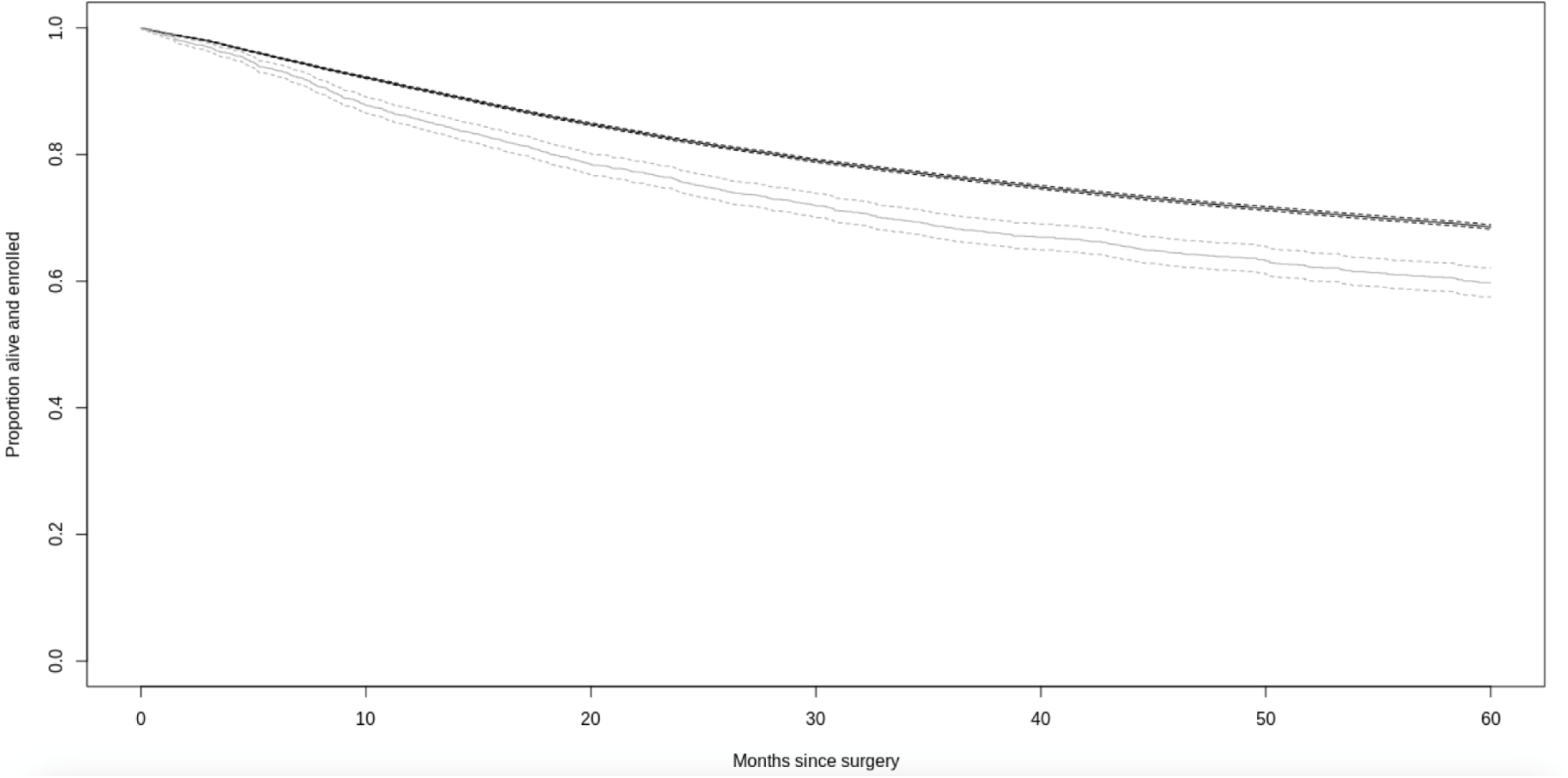
Crude survival in the five years following cancer surgery by HIV status at surgery. Notes: Survival for PWH is in light grey, and survival for PWoH is in black.

**Figure 3.**
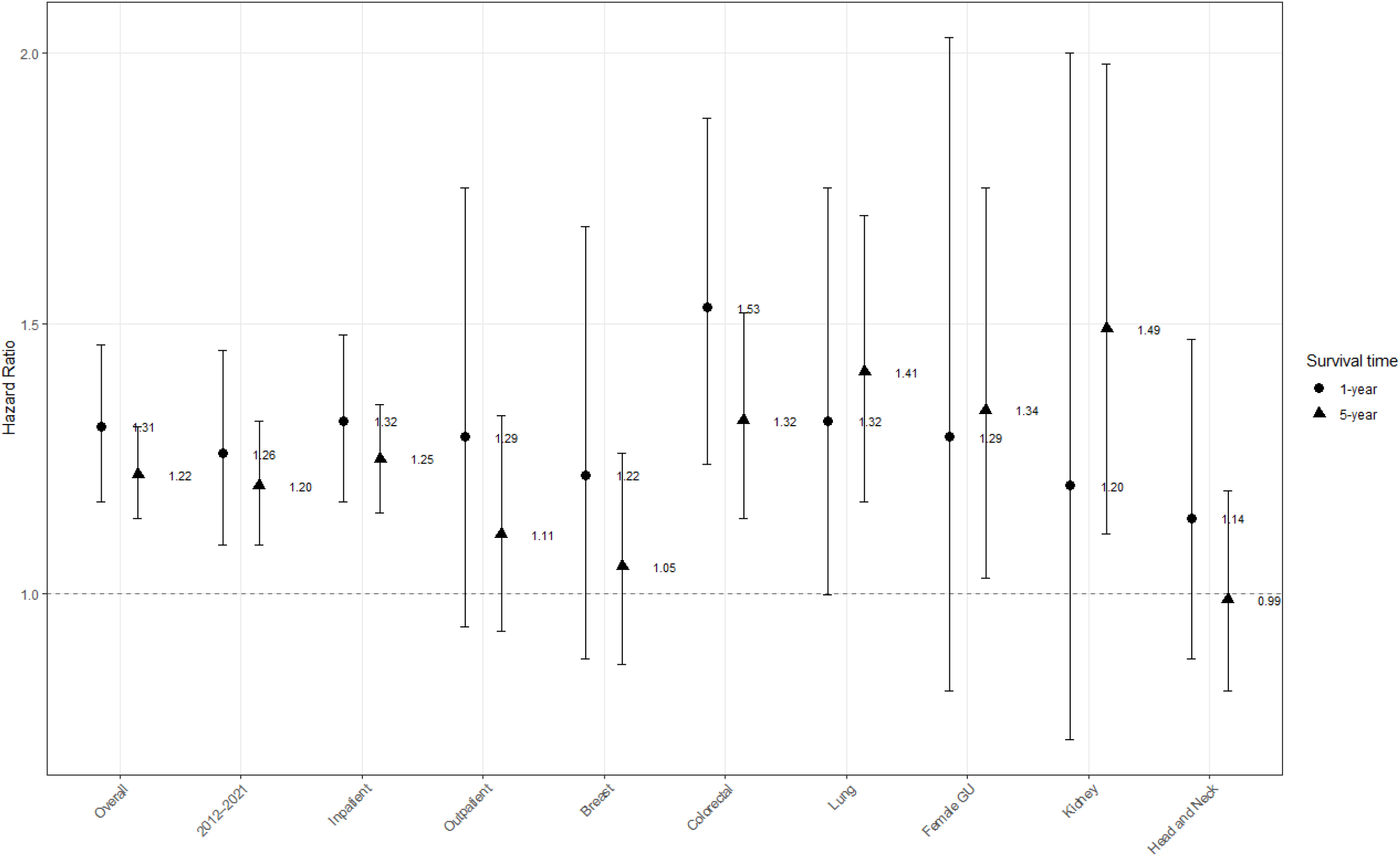
Adjusted hazard ratios for Cox proportional hazards models of 1- and 5-year survival overall and by subgroup. Models adjusts for age, sex, race, state, year of cancer surgery, cancer type, comorbidity count, receipt of additional cancer treatment with separate indicators for oral chemotherapy or immunotherapy, infusion chemotherapy, or radiation, surgical risk, and metastasis

## Discussion

In this large-scale retrospective cohort study of short-term utilization, complications, and short- and long-term mortality following an initial cancer surgery among Medicaid beneficiaries, we observed elevated utilization among PWH after cancer treatment, but no significant differences in surgical complications like SSI or 30-day mortality. These findings were consistent when restricting by cancer type and surgical setting and adjusting for surgical risk, receipt of other cancer therapy, and metastasis at the time of surgery. Although 90-day mortality was significantly different overall, there were no significant differences when limiting to the treat-all ART era. These findings taken together align with prior literature among PWH undergoing lung cancer resection at the VA^12^ and indicate that PWH are not at significantly elevated risk of immediate short-term complications of cancer surgery and should be offered standard of care in cancer treatment.^26^

While we did not observe elevated short-term mortality or SSI, we did find increased LOS, days in hospital, and ED visits among PWH, which replicates prior literature indicating that PWH have higher rates of ED utilization, hospital admission, and hospital readmission broadly,^15–17,27^ and extends the results to the post-cancer surgery setting. These differences may reflect increased comorbidity management burden and care complexity rather than complications directly related to surgery among PWH.^28,29^ While numerous factors may drive differences in utilization, interventions such as post-discharge referral and enhanced care coordination may help reduce readmissions and acute care utilization among PWH, though additional research is needed to identify specific risk factors.^30^

We also observed that PWH had significantly higher long-term mortality than PWoH following a first observed cancer surgery, overall and in the treat-all ART era. Prior literature has also found increased 5-year mortality after cancer diagnosis accounting for stage among PWH.^8,9,31–33^ Similar differences were found in our study but limited to inpatient surgeries only. Outpatient surgeries did not have statistically significant mortality differences, which likely reflects underlying differences in stage at diagnosis and cancer type determining site of surgery. When examining specific cancers, colorectal cancer and lung cancer had the largest mortality differences for PWH. Although we adjusted for surgical risk and presence of metastatic disease, residual differences in surgical procedure and stage at diagnosis could contribute to mortality differences. While we accounted for additional lines of cancer therapy at the time of surgery, we did not characterize the full course of cancer treatment and prior literature has suggested that PWH may be less likely to receive standard cancer treatment.^34,35^

Long-term survival differences may also be impacted by increased risk of non-cancer mortality among PWH. We previously found a significant and prolonged CD4 cell count decline among PWH receiving chemotherapy or radiation that was associated with increased mortality risk among PWH diagnosed with cancer from 1997 - 2016.^36^ We also reported an increase in the incidence of new AIDS-defining illness after cancer-treatment among PWH.^37^ Further information on receipt of additional cancer treatment, the impact of cancer treatment on risk of AIDS-defining illness or HIV progression, and stage at diagnosis is needed to determine drivers of these mortality differences.

There are several limitations to consider when interpreting our results. First, stage at cancer diagnosis is not captured in claims data. To mitigate this limitation, we used the Charlson Comorbidity Index definition for cancer metastasis^22^ to capture advanced cancers at the time of surgery. This approach has a positive predictive value ranging from 75% to 85% depending on cancer type.^38^ The long-term mortality differences by HIV status may reflect differences in stage at cancer diagnosis that we previously observed using HIV clinical cohort data linked to cancer registry information.^35^ Second, to account for differences in underlying risk of SSI, readmission, and short-term mortality across a wide range of surgical procedures with small counts for most individual codes, we categorized individual procedure codes into surgical risk categories, which may not fully capture differences risk by procedure type. It is likely that there is some residual confounding due to this classification approach. Despite these limitations, we were able to leverage the strengths of Medicaid data to be able to capture cancer surgery outcomes for a large proportion of PWH in the United States, as Medicaid insures 40% of PWH in the US and compare outcomes with a socioeconomically similar comparison group.^19^ The large sample size enabled us to examine differences by cancer type, surgical setting, and time period.

Among a large cohort of Medicaid beneficiaries undergoing cancer surgery, we observed higher acute care utilization for PWH but no increased short-term surgical complications, which supports the current guidelines to provide standard cancer care for PWH. However, we observed higher long-term mortality following surgery for PWH. Future research should focus on identifying drivers of this disparity, including stage at diagnosis, differences in receipt of subsequent cancer therapies in accordance with the guidelines, and the impact of cancer treatment on HIV disease progression. Linkage of administrative claims to cancer registry could help further this research. Finally, evidence on potential interventions after hospitalization that may both reduce utilization and improve long-term survival are needed.

## Data Availability

The data that support the findings of this study are available from the Centers for Medicare & Medicaid Services (CMS) and administered by ResDAC. but restrictions apply to the availability of these data, which were used under license for the current study, and so are not publicly available. Investigators may reuse these data if they independently meet CMS requirements and obtain both CMS reuse approval and permission from the study NIH program officer. Investigators may contact.

## Availability of data and materials

The data that support the findings of this study are available from the Centers for Medicare & Medicaid Services (CMS) and administered by ResDAC. but restrictions apply to the availability of these data, which were used under license for the current study, and so are not publicly available. Investigators may reuse these data if they independently meet CMS requirements and obtain both CMS reuse approval and permission from the study’s NIH program officer. Investigators may contact.

