## Supplemental Tables and Figures for "Differences in utilization, complications, and mortality after cancer surgery by HIV status among Medicaid beneficiaries from 2001-2021"

**Table S1. List of ICD-9-CM and ICD-10-CM diagnosis codes used in the study.**

| Variable | ICD-9-CM Codes[a] | ICD-10-CM Codes[a] |
|---|---|---|
| **HIV** | 042-044, 079.53, 795.71, V08 | B20, B97.35, R75, Z21 |
| **Cancers** | | |
| Anal | 154.2, 154.3 | C21.0, C21.1, C21.2 |
| Bladder | 188.X | C67.X |
| Breast | 174.X, 175.X | C50.X |
| Colorectal | 153.X, 154.0, 154.1 | C18.X, C19.X, C20.X |
| Female Genitourinary | 179.X, 180.X, 182.X, 184.X | C51.X, C52.X, C53.X, C54.X, C55.X |
| Gastroesophageal | 150.X, 151.X | C15.X, C16.X |
| Head and Neck | 140.X-149.X, 160.X, 161.X | C01.X-C05.X, C09.X, C10.X |
| Kidney | 189.X | C64.X, C65.X, C66.X, C68.X |
| Liver | 155.X | C22.X |
| Lung | 162.2, 162.3, 162.4, 162.5, 162.8, 163.9 | C34.X |
| Ovarian | 183.X | C56.X, C57.X |
| Pancreatic | 157.X | C60.X |
| **Comorbidities** | | |
| Myocardial infarction | 410.X, 412.X | I21.X, I22.X, I25.2 |
| Congestive heart failure | 398.91, 402.01, 402.11, 402.91, 404.01, 404.03, 404.11, 404.13, 404.91, 404.93, 425.4-425.9, 428.X | I09.8, I11.0, I13.0, I13.2, I25.5, I42.0, I42.5 – I42.9, I43.X, I50.X, P29.0 |
| Peripheral vascular disease | 093.0, 437.3, 440.X, 441.X, 443.1-443.9, 447.1, 557.1, 557.9, V43.4 | I70.X, I71.X, I73.1, II73.8, I73.9, I77.1, I79.0, I79.2, K55.1, K55.8, K55.9, Z95.8, Z95.9 |
| Cerebrovascular disease | 362.34, 430.X-438.X | G45.X, G46.X, H34.0, I60.X – I 69.X |
| Diabetes with or without chronic complication | 250.0-250.9 | E10.X, E11.X, E12.X, E13.X, E14.X |

| | | |
|---|---|---|
| Dementia | 290.X, 294.1, 331.2 | F00.X – F03.X, F05.1, G30.X, G31.1 |
| Chronic pulmonary disease | 416.8, 416.9, 490.X – 505.X, 506.4, 508.1, 508.8 | I27.8, I27.9, J40.X – J47.X, J60.X – J67.X, J68.4, J70.1, J70.3 |
| Rheumatic disease | 446.5, 710.0 – 710.4, 714.0 – 714.2, 714.8, 725.X | M05.X, M06.X, M31.5, M32.X – M34.X, M35.1, M35.3, M36.0 |
| Mild or moderate liver disease | 070.22, 070.23, 070.32, 070.33, 070.44, 070.54, 070.6, 070.9, 456.0 – 456.2, 570.X, 571.X, 572.2-572.8, 573.3, 573.4, 573.8, 573.9, V42.7 | B18.X, , I85.0, I85.9, I86.4, I98.2, K70.0 – K70.4, K70.9, K71.1, K71.3 – K71.5, K71.7, K72.1, K72.9, K73.X, K74.X, K76.0, K76.2 – K76.9, Z94.4 |
| Hemiplegia or paraplegia | 334.1, 342.X, 343.X, 344.0 – 344.6, 344.9 | G04.1, G11.4, G80.1, G80.2, G81.X, G82.X, G83.0 – G83.4, G83.9 |
| Renal disease | 403.01, 403.11, 403.91, 404.02, 404.03, 404.12, 404.13, 404.92, 404.93, 582.X, 582.0 – 583.7, 585.X, 586.X, 588.0, V42.0 V45.1, V56.X | I12.0, I13.1, N03.2 – N03.7, N05.2 – N05.7, N18.X, N19.X, N25.0, Z49.0 – Z49.2, Z94.0, Z99.2 |
| Peptic ulcer disease | 531.X – 534.X | K25.X – K28.X |
| Metastatic disease | 196.X – 199.X | C77.X – C80.X |

**[a] A code ending with ".X" indicates a wildcard. Any number could appear after the decimal place.**

**Table S2. List of ICD-9-PCS, ICD-10-PCS, and CPT procedure codes used to identify cancer surgeries.**

| Cancer surgery site | ICD-9-PCS Codes[a] | ICD-10-PCS Codes[a] | CPT Codes |
|---|---|---|---|
| Anal | 45.95, 48.42, 48.49-48.52, 48.59, 48.62-48.63, 48.69, 49.6, 57.71, 68.8 | 0D1B0ZQ, 0D1B4ZQ, 0D1B8ZQ, 0DBP0ZZ, 0DBP4ZZ, 0DBQ0ZZ, 0DBQ3ZZ, 0DBQ4ZZ, 0DBQ7ZZ, 0DBQ8ZZ, 0DBQXZZ, 0DBR0ZZ, 0DBR3ZZ, 0DBR4ZZ, 0DTP0ZZ, 0DTP4ZZ, 0DTP7ZZ, 0DTP8ZZ, 0DTQ0ZZ, 0DTQ4ZZ, 0DTQ7ZZ, 0DTQ8ZZ, 0DTR0ZZ, 0DTR4ZZ, 0TTB0ZZ, 0TTB4ZZ, 0TTB7ZZ, 0TTB8ZZ, 0TTD0ZZ, 0TTD4ZZ, 0TTD7ZZ, 0TTD8ZZ, 0UT20ZZ, 0UT70ZZ, 0UT90ZZ, 0UTC0ZZ, 0UTG0ZZ, 0VT00ZZ, 0VT04ZZ, 0VT07ZZ, 0VT08ZZ, 0VT30ZZ, 0VT34ZZ | 11970-11971, 19120, 19125-19126, 19294, 19301-19307, 19340, 19342, 19350, 19357, 19361, 19364, 19366-19367, 19369, 19380, 21601-21603, 28525, 0546T, 0581T |
| Bladder | 57.71, 57.79, 57.87, 68.8 | 0TRB07Z, 0TRB0JZ, 0TRB0KZ, 0TRB47Z, 0TRB4JZ, 0TRB4KZ, 0TRB77Z, 0TRB7JZ, 0TRB7KZ, 0TRB87Z, 0TRB8JZ, 0TRB8KZ, 0TTB0ZZ, 0TTB4ZZ, 0TTB7ZZ, 0TTB8ZZ, 0TTD0ZZ, 0TTD4ZZ, 0TTD7ZZ, 0TTD8ZZ, 0TUB07Z, 0TUB0JZ, 0TUB0KZ, 0TUB47Z, 0TUB4JZ, 0TUB4KZ, 0TUB77Z, 0TUB7JZ, 0TUB7KZ, 0TUB87Z, 0TUB8JZ, 0TUB8KZ, 0UT20ZZ, 0UT70ZZ, 0UT90ZZ, 0UTC0ZZ, 0UTG0ZZ, 0VT00ZZ, 0VT04ZZ, 0VT07ZZ, 0VT08ZZ, 0VT30ZZ, 0VT34ZZ | 58548, 58550, 58552, 58950, 58953 |
| Breast | 85.20-85.23, 85.41-85.48, 85.53-85.54, 85.70, 85.79, 85.83-85.84, 85.93-85.96 | 0H0T0JZ, 0H0T0KZ, 0H0T3JZ, 0H0T3KZ, 0H0U0JZ, 0H0U0KZ, 0H0U3JZ, 0H0U3KZ, 0H0V07Z, 0H0V0JZ, 0H0V0KZ, 0H0V37Z, 0H0V3JZ, 0H0V3KZ, 0H5T0ZZ, 0H5T3ZZ, 0H5T7ZZ, 0H5T8ZZ, 0H5TXZZ, 0H5U0ZZ, 0H5U3ZZ, 0H5U7ZZ, 0H5U8ZZ, 0H5UXZZ, 0H5V0ZZ, 0H5V3ZZ, 0H5V7ZZ, 0H5V8ZZ, 0H5VXZZ, 0HBT0ZZ, 0HBT3ZZ, 0HBT7ZZ, 0HBT8ZZ, 0HBTXZZ, 0HBU0ZZ, 0HBU3ZZ, 0HBU7ZZ, 0HBU8ZZ, 0HBUXZZ, 0HBV0ZZ, 0HBV3ZZ, 0HBV7ZZ, 0HBV8ZZ, 0HBVXZZ, 0HDT0ZZ, 0HDU0ZZ, 0HDV0ZZ, 0HDY0ZZ, 0HHT0NZ, 0HHT3NZ, 0HHT7NZ, 0HHT8NZ, 0HHU0NZ, 0HHU3NZ, 0HHU7NZ, 0HHU8NZ, 0HHV0NZ, 0HHV3NZ, 0HHV7NZ, 0HHV8NZ, 0HHW0NZ, 0HHW3NZ, 0HHW7NZ, 0HHW8NZ, 0HHX0NZ, 0HHX3NZ, 0HHX7NZ, 0HHX8NZ, 0HPT0JZ, 0HPT0NZ, 0HPT3JZ, 0HPT3NZ, 0HPU0JZ, 0HPU0NZ, 0HPU3JZ, 0HPU3NZ, 0HRT07Z, 0HRT0JZ, 0HRT0KZ, 0HRT3JZ, 0HRT3KZ, 0HRTX7Z, 0HRTXKZ, 0HRU07Z, 0HRU0JZ, 0HRU0KZ, 0HRU3JZ, 0HRU3KZ, 0HRUX7Z, 0HRUXKZ, 0HRVX7Z, 0HRVXKZ, 0HTT0ZZ, 0HTU0ZZ, 0HTV0ZZ, 0HUT0JZ, 0HUT0KZ, 0HUT3JZ, 0HUT3KZ, 0HUU0JZ, 0HUU0KZ, 0HUU3JZ, 0HUU3KZ, 0HUV0JZ, 0HUV3JZ, 0HWT0JZ, 0HWT3JZ, 0HWU0JZ, 0HWU3JZ, 0HX5XZZ, 0KTH0ZZ, 0KTJ0ZZ | 43122, 43276-43277 |
| Colorectal | 65.49, 65.51-65.53, 65.61-65.62, 68.31, 68.39, 68.41, 68.49, 68.51, 68.61, 68.69, 68.71, 68.79, 68.8-68.9 | 0DXE0Z7, 0DXE4Z7, 0TTB0ZZ, 0TTD0ZZ, 0U508ZZ, 0U518ZZ, 0U528ZZ, 0U548ZZ, 0UT00ZZ, 0UT07ZZ, 0UT08ZZ, 0UT0FZZ, 0UT10ZZ, 0UT17ZZ, 0UT18ZZ, 0UT1FZZ, 0UT20ZZ, 0UT24ZZ, 0UT27ZZ, 0UT28ZZ, 0UT2FZZ, 0UT40ZZ, 0UT44ZZ, 0UT70ZZ, 0UT90ZL, 0UT90ZZ, 0UT94ZL, 0UT94ZZ, 0UT97ZL, 0UT97ZZ, 0UT98ZL, 0UT98ZZ, 0UT9FZL, 0UT9FZZ, 0UTC0ZZ, 0UTC4ZZ, 0UTC7ZZ, 0UTC8ZZ, 0UTG0ZZ | 21299, 21554, 21558, 31420, 31599, 41120, 42844, 42890, 42950 |
| Female Genitourinary | 42.40-42.42, 42.51-42.56, 42.58-42.59, 42.61-42.65, 42.69, 43.42, 43.81-43.82, 43.89, 43.91, 43.99, 46.39 | 06L27CZ, 06L27DZ, 06L27ZZ, 06L28CZ, 06L28DZ, 06L28ZZ, 0D15074, 0D15076, 0D15079, 0D1507A, 0D1507B, 0D150J4, 0D150J6, 0D150J9, 0D150JA, 0D150JB, 0D150K4, 0D150K6, 0D150K9, 0D150KA, 0D150KB, 0D150Z4, 0D150Z6, 0D150Z9, 0D150ZA, 0D150ZB, 0D153J4, 0D15474, 0D15476, 0D15479, 0D1547A, 0D1547B, 0D154J4, 0D154J6, 0D154J9, 0D154JA, 0D154JB, 0D154K4, 0D154K6, 0D154K9, 0D154KA, 0D154KB, 0D154Z4, 0D154Z6, 0D154Z9, 0D154ZA, 0D154ZB, 0D15876, 0D15879, 0D1587A, 0D1587B, 0D158J6, 0D158J9, 0D158JA, 0D158JB, 0D158K6, 0D158K9, 0D158KA, 0D158KB, 0D158Z6, 0D158Z9, 0D158ZA, 0D158ZB, 0D188KM, 0D188KN, 0D188KP, 0D188KQ, 0D188Z4, 0D188Z8, 0D188ZH, 0D188ZK, 0D188ZL, 0D188ZM, 0D188ZN, 0D188ZP, 0D188ZQ, 0D1E074, 0D1E07E, 0D1E07P, 0D1E0J4, 0D1E0JE, 0D1E0JP, 0D1E0K4, 0D1E0KE, 0D1E0KP, 0D1E0Z4, 0D1E0ZE, 0D1E0ZP, 0D1E474, 0D1E47E, 0D1E47P, 0D1E4J4, 0D1E4JE, 0D1E4JP, 0D1E4K4, 0D1E4KE, 0D1E4KP, 0D1E4Z4, 0D1E4ZE, 0D1E4ZP, 0D1E874, 0D1E87E, 0D1E87P, 0D1E8J4, 0D1E8JE, 0D1E8JP, 0D1E8K4, 0D1E8KE, 0D1E8KP, 0D1E8Z4, 0D1E8ZE, 0D1E8ZP, 0DB50ZZ, 0DB53ZZ, 0DB57ZZ, 0DB60ZZ, 0DB63ZZ, | 50220, 50225, 50230, 50234, 50236, 50240, 50543, 50545-50546, 50548 |

| | | | |
|---|---|---|---|
| | | 0DB64Z3, 0DB64ZZ, 0DB67ZZ, 0DB68ZZ, 0DH80UZ, 0DH83UZ, 0DH84UZ, 0DH87UZ, 0DH88UZ, 0DH90UZ, 0DH93UZ, 0DH94UZ, 0DH97UZ, 0DH98UZ, 0DHA0UZ, 0DHA7UZ, 0DT50ZZ, 0DT54ZZ, 0DT57ZZ, 0DT58ZZ, 0DT60ZZ, 0DT64ZZ, 0DT67ZZ, 0DT68ZZ, 0DX60Z5, 0DX64Z5, 0DX80Z5, 0DX84Z5, 0DXE0Z5, 0DXE0Z7, 0DXE4Z5, 0DXE4Z7 | |
| Gastroesophageal | 21.4-21.5, 21.69, 25.2-25.4, 27.42, 27.57, 29.33, 30.1, 30.21-30.22, 30.29, 30.3-30.4, 40.40, 40.42, 76.31, 76.43, 76.45, 76.91 | 07T10ZZ, 07T14ZZ, 07T20ZZ, 07T24ZZ, 09BL0ZZ, 09BL3ZZ, 09BL4ZZ, 09BL7ZZ, 09BL8ZZ, 09BM0ZZ, 09BM3ZZ, 09BM4ZZ, 09BN0ZZ, 09BN3ZZ, 09BN4ZZ, 09BN7ZZ, 09BN8ZZ, 09TK0ZZ, 09TK4ZZ, 09TKXZZ, 09TL0ZZ, 09TL4ZZ, 09TL7ZZ, 09TL8ZZ, 09TM0ZZ, 09TM4ZZ, 09TN0ZZ, 09TN4ZZ, 09TN7ZZ, 09TN8ZZ, 0C5R0ZZ, 0C5R3ZZ, 0C5R4ZZ, 0C5R7ZZ, 0C5R8ZZ, 0CB00ZZ, 0CB03ZZ, 0CB0XZZ, 0CB10ZZ, 0CB13ZZ, 0CB1XZZ, 0CB70ZZ, 0CB73ZZ, 0CB7XZZ, 0CBM0ZZ, 0CBM3ZZ, 0CBM4ZZ, 0CBM7ZZ, 0CBM8ZZ, 0CBR0ZZ, 0CBR3ZZ, 0CBR4ZZ, 0CBR7ZZ, 0CBR8ZZ, 0CBS0ZZ, 0CBS3ZZ, 0CBS4ZZ, 0CBS7ZZ, 0CBS8ZZ, 0CBT0ZZ, 0CBT3ZZ, 0CBT4ZZ, 0CBT7ZZ, 0CBT8ZZ, 0CBV0ZZ, 0CBV3ZZ, 0CBV4ZZ, 0CBV7ZZ, 0CBV8ZZ, 0CT70ZZ, 0CT7XZZ, 0CTM0ZZ, 0CTM4ZZ, 0CTM7ZZ, 0CTM8ZZ, 0CTR0ZZ, 0CTR4ZZ, 0CTR7ZZ, 0CTR8ZZ, 0CTS0ZZ, 0CTS4ZZ, 0CTS7ZZ, 0CTS8ZZ, 0CTT0ZZ, 0CTT4ZZ, 0CTT7ZZ, 0CTT8ZZ, 0CTV0ZZ, 0CTV4ZZ, 0CTV7ZZ, 0CTV8ZZ, 0CX00ZZ, 0CX0XZZ, 0CX10ZZ, 0CX1XZZ, 0CX40ZZ, 0CX4XZZ, 0CX50ZZ, 0CX5XZZ, 0CX60ZZ, 0CX6XZZ, 0NBT0ZZ, 0NBT3ZZ, 0NBT4ZZ, 0NBV0ZZ, 0NBV3ZZ, 0NBV4ZZ, 0NQT0ZZ, 0NQT3ZZ, 0NQT4ZZ, 0NQTXZZ, 0NQV0ZZ, 0NQV3ZZ, 0NQV4ZZ, 0NQVXZZ, 0NRC07Z, 0NRC0KZ, 0NRC37Z, 0NRC3KZ, 0NRC47Z, 0NRC4KZ, 0NRD07Z, 0NRD0KZ, 0NRD37Z, 0NRD3KZ, 0NRD47Z, 0NRD4KZ, 0NRF07Z, 0NRF0KZ, 0NRF37Z, 0NRF3KZ, 0NRF47Z, 0NRF4KZ, 0NRG07Z, 0NRG0KZ, 0NRG37Z, 0NRG3KZ, 0NRG47Z, 0NRG4KZ, 0NRH07Z, 0NRH0KZ, 0NRH37Z, 0NRH3KZ, 0NRH47Z, 0NRH4KZ, 0NRJ07Z, 0NRJ0KZ, 0NRJ37Z, 0NRJ3KZ, 0NRJ47Z, 0NRJ4KZ, 0NRK07Z, 0NRK0KZ, 0NRK37Z, 0NRK3KZ, 0NRK47Z, 0NRK4KZ, 0NRL07Z, 0NRL0KZ, 0NRL37Z, 0NRL3KZ, 0NRL47Z, 0NRL4KZ, 0NRM07Z, 0NRM0KZ, 0NRM37Z, 0NRM3KZ, 0NRM47Z, 0NRM4KZ, 0NRN07Z, 0NRN0KZ, 0NRN37Z, 0NRN3KZ, 0NRN47Z, 0NRN4KZ, 0NRP0KZ, 0NRP3KZ, 0NRP4KZ, 0NRQ0KZ, 0NRQ3KZ, 0NRQ4KZ, 0NTC0ZZ, 0NTD0ZZ, 0NTF0ZZ, 0NTG0ZZ, 0NTH0ZZ, 0NTJ0ZZ, 0NTK0ZZ, 0NTL0ZZ, 0NTM0ZZ, 0NTN0ZZ, 0NTP0ZZ, 0NTQ0ZZ, 0NTR0ZZ, 0NTS0ZZ, 0NUC07Z, 0NUC0KZ, 0NUC37Z, 0NUC3KZ, 0NUC47Z, 0NUC4KZ, 0NUD07Z, 0NUD0KZ, 0NUD37Z, 0NUD3KZ, 0NUD47Z, 0NUD4KZ, 0NUF07Z, 0NUF0KZ, 0NUF37Z, 0NUF3KZ, 0NUF47Z, 0NUF4KZ, 0NUG07Z, 0NUG0KZ, 0NUG37Z, 0NUG3KZ, 0NUG47Z, 0NUG4KZ, 0NUH07Z, 0NUH0KZ, 0NUH37Z, 0NUH3KZ, 0NUH47Z, 0NUH4KZ, 0NUJ07Z, 0NUJ0KZ, 0NUJ37Z, 0NUJ3KZ, 0NUJ47Z, 0NUJ4KZ, 0NUK07Z, 0NUK0KZ, 0NUK37Z, 0NUK3KZ, 0NUK47Z, 0NUK4KZ, 0NUL07Z, 0NUL0KZ, 0NUL37Z, 0NUL3KZ, 0NUL47Z, 0NUL4KZ, 0NUM07Z, 0NUM0KZ, 0NUM37Z, 0NUM3KZ, 0NUM47Z, 0NUM4KZ, 0NUN07Z, 0NUN0KZ, 0NUN37Z, 0NUN3KZ, 0NUN47Z, 0NUN4KZ, 0NUP07Z, 0NUP0KZ, 0NUP37Z, 0NUP3KZ, 0NUP47Z, 0NUP4KZ, 0NUQ07Z, 0NUQ0KZ, 0NUQ37Z, 0NUQ3KZ, 0NUQ47Z, 0NUQ4KZ, 0NUT07Z, 0NUT0JZ, 0NUT0KZ, 0NUT37Z, 0NUT3JZ, 0NUT3KZ, 0NUT47Z, 0NUT4JZ, 0NUT4KZ, 0NUV07Z, 0NUV0JZ, 0NUV0KZ, 0NUV37Z, 0NUV3JZ, 0NUV3KZ, 0NUV47Z, 0NUV4JZ, 0NUV4KZ, 0RTC0ZZ, 0RTD0ZZ, 0WC40ZZ, 0WC43ZZ, 0WC44ZZ, 0WC50ZZ, 0WC53ZZ, 0WC54ZZ | |
| Head and Neck | 55.5, 55.51-55.52 | 0TT00ZZ, 0TT04ZZ, 0TT10ZZ, 0TT14ZZ | |
| Kidney | 50.22, 50.3-50.4, 50.59, 50.99 | 0F500ZF, 0F503ZF, 0F504ZF, 0F510ZF, 0F513ZF, 0F514ZF, 0F520ZF, 0F523ZF, 0F524ZF, 0FB00ZZ, 0FB03ZZ, 0FB04ZZ, 0FQ00ZZ, 0FQ03ZZ, 0FQ04ZZ, 0FT00ZZ, 0FT04ZZ, 0FT10ZZ, 0FT14ZZ, 0FT20ZZ, 0FT24ZZ, 0FY00Z0, 0FY00Z1, 0FY00Z2 | 58548, 58550, 58552, 58950, 58953 |
| Liver | 32.20-32.30, 32.39, 32.41, 32.49-32.50, 32.59 | 0B538ZZ, 0B548ZZ, 0B558ZZ, 0B568ZZ, 0B578ZZ, 0B588ZZ, 0B598ZZ, 0B5B8ZZ, 0B5K0ZZ, 0B5K3ZZ, 0B5K4ZZ, 0B5K7ZZ, 0B5K8ZZ, 0B5L0ZZ, 0B5L3ZZ, 0B5L4ZZ, 0B5L7ZZ, 0B5L8ZZ, 0B5M0ZZ, 0B5M3ZZ, 0B5M4ZZ, 0B5M7ZZ, 0B5M8ZZ, 0BBC4ZZ, 0BBD4ZZ, 0BBF4ZZ, 0BBG4ZZ, 0BBH4ZZ, 0BBJ4ZZ, 0BBK0ZZ, 0BBK3ZZ, 0BBK4ZZ, 0BBK7ZZ, 0BBK8ZZ, 0BBL0ZZ, 0BBL3ZZ, 0BBL4ZZ, 0BBL7ZZ, 0BBL8ZZ, 0BBM0ZZ, 0BBM3ZZ, 0BBM4ZZ, 0BBM7ZZ, 0BBM8ZZ, 0BQK0ZZ, | 48999 |

| | | | |
|---|---|---|---|
| | | 0BQK3ZZ, 0BQK4ZZ, 0BQK7ZZ, 0BQK8ZZ, 0BQL0ZZ, 0BQL3ZZ, 0BQL4ZZ, 0BQL7ZZ, 0BQL8ZZ, 0BQM0ZZ, 0BQM3ZZ, 0BQM4ZZ, 0BQM7ZZ, 0BQM8ZZ, 0BTC0ZZ, 0BTC4ZZ, 0BTD0ZZ, 0BTD4ZZ, 0BTF0ZZ, 0BTF4ZZ, 0BTG0ZZ, 0BTG4ZZ, 0BTH4ZZ, 0BTJ0ZZ, 0BTJ4ZZ, 0BTK0ZZ, 0BTK4ZZ, 0BTL0ZZ, 0BTL4ZZ, 0BTM0ZZ, 0BTM4ZZ | |
| Lung | 65.49, 65.51-65.53, 65.61-65.62, 68.31, 68.39, 68.41, 68.49, 68.51, 68.61, 68.69, 68.71, 68.79, 68.8-68.9 | 0TTB0ZZ, 0TTD0ZZ, 0U508ZZ, 0U518ZZ, 0U528ZZ, 0U548ZZ, 0UT00ZZ, 0UT07ZZ, 0UT08ZZ, 0UT0FZZ, 0UT10ZZ, 0UT17ZZ, 0UT18ZZ, 0UT1FZZ, 0UT20ZZ, 0UT24ZZ, 0UT27ZZ, 0UT28ZZ, 0UT2FZZ, 0UT40ZZ, 0UT44ZZ, 0UT70ZZ, 0UT90ZL, 0UT90ZZ, 0UT94ZL, 0UT94ZZ, 0UT97ZL, 0UT97ZZ, 0UT98ZL, 0UT98ZZ, 0UT9FZL, 0UT9FZZ, 0UTC0ZZ, 0UTC4ZZ, 0UTC7ZZ, 0UTC8ZZ, 0UTG0ZZ | 52601, 52648, 53854, 55845, 55874 |
| Ovarian | 52.52-52.53, 52.6-52.7 | 0F5G0ZF, 0F5G3ZF, 0F5G4ZF, 0FBG0ZZ, 0FBG3ZZ, 0FBG4ZZ, 0FTG0ZZ, 0FTG4ZZ | 44120, 44140, 44204, 44207, 45123, 47100, 47120, 47370, 47382 |
| Pancreatic | 57.71, 60.21, 60.29, 60.3-60.5, 60.69 | 0TTB0ZZ, 0TTB4ZZ, 0TTB7ZZ, 0TTB8ZZ, 0TTD0ZZ, 0TTD4ZZ, 0TTD7ZZ, 0TTD8ZZ, 0V507ZZ, 0V508ZZ, 0VB07ZZ, 0VB08ZZ, 0VT00ZZ, 0VT04ZZ, 0VT07ZZ, 0VT08ZZ, 0VT30ZZ, 0VT34ZZ, XV508A4 | 11970-11971, 19120, 19125-19126, 19294, 19301-19307, 19340, 19342, 19350, 19357, 19361, 19364, 19366-19367, 19369, 19380, 21601-21603, 28525, 0546T, 0581T |

**[a] A code ending with ".X" indicates a wildcard. Any number could appear after the decimal place.**

**Table S3. List of HCPCS, NDC, ICD-9-PCS, and ICD-10-PCS codes used to identify other cancer treatment modalities.**

| **Variable** | **Codes** |
|---|---|
| Infusion Chemotherapy | **HCPCS:** A9513, A9543, A9545, A9590, C9021, C9024, C9025, C9027, C9028, C9030, C9031, C9042, C9044, C9045, C9049, C9062, C9065, C9066, C9069, C9070, C9073, C9131, C9287, C9289, C9292, C9295, C9296, C9297, C9408, C9442, C9449, C9453, C9455, C9467, C9472, C9474, C9475, C9476, C9477, C9480, C9483, C9485, C9491, C9492, J0202, J0594, J0894, J1930, J2353, J2354, J2860, J7504, J7511, J8510, J8520, J8521, J8530, J8560, J8562, J8565, J8600, J8700, J8705, J8999, J9000, J9001, J9002, J9010, J9015, J9017, J9019, J9020, J9022, J9023, J9025, J9027, J9032, J9033, J9034, J9035, J9036, J9039, J9040, J9041, J9042, J9043, J9044, J9045, J9047, J9050, J9055, J9057, J9060, J9062, J9065, J9070, J9080, J9090, J9091, J9092, J9093, J9094, J9095, J9096, J9097, J9098, J9100, J9118, J9119, J9120, J9130, J9140, J9144, J9145, J9150, J9151, J9153, J9160, J9170, J9171, J9173, J9176, J9177, J9178, J9179, J9181, J9182, J9185, J9190, J9198, J9199, J9200, J9201, J9203, J9205, J9206, J9207, J9208, J9211, J9216, J9223, J9227, J9228, J9229, J9230, J9245, J9246, J9261, J9262, J9263, J9264, J9265, J9266, J9267, J9268, J9269, J9271, J9285, J9293, J9295, J9299, J9300, J9301, J9302, J9303, J9304, J9305, J9306, J9307, J9308, J9309, J9310, J9311, J9312, J9313, J9315, J9316, J9317, J9320, J9325, J9328, J9330, J9340, J9350, J9351, J9352, J9354, J9355, J9356, J9357, J9358, J9360, J9370, J9371, J9375, J9380, J9390, J9395, J9400, J9999, Q2017, Q2040, Q2041, Q2042, Q2043, Q2048, Q2049, Q2050, Q5107, Q5112, Q5113, Q5114, Q5115, Q5116, Q5117, Q5118, Q5119, Q9979, WW002, WW003, WW004, WW005, WW006, WW007, WW008, WW009, WW020, WW030, WW031, WW032, WW080, WW081, WW089, WW090, WW091, WW093, WW094, WW096, WW140 |
| Oral Chemotherapy | **NDC**: 100190925-100190927, 100190929, 100190935-100190939, 100190942-100190945, 100190955-100190957, 100190988-100190990, 101390063, 101390321, 101911989, 108850001, 113990005, 1151675, 125160592, 125168003-125168004, 127140161-127140163, 127140905, 128540549, 128540551, 128540803-128540805, 130132, 132576, 132586, 132596, 139250523, 1439202-1439203, 1439217-1439219, 1439240-1439241, 1439245, 1439270, 1439275, 1439277, 1439306-1439309, 1439369-1439372, 1439376, 1439504-1439505, 1439510-1439512, 1439530-1439531, 1439546-1439551, 1439565, 1439583, 1439597, 1439606, 1439701-1439702, 1439871, 147890600, 150502-150506, 150540043, 150540060, 150540090, 150540120, 150541060, 150541090, 150541120, 150556-150557, 151910-151911, 153012, 153030-153032, 153061-153062, 153072, 153075, 153080, 153084, 153091, 153095, 153210-153216, 153380237, 153380255, 153380335, 153380866-153380868, 153404, 153475-153476, 153479, 163640048-163640051, 163640072-163640073, 163640114, 163640129-163640130, 163640231, 163640243, 163640267, 163640276, 163640424, 167140027, 167140118, 167140131, 167140465, 167140467-167140468, 167140500, 167140704-167140705, 167140725-167140728, 167140742, 167140749, 167140777, 167140856-167140859, 167140909, 167140927-167140928, 167140930, 167140963, 167290048-167290051, 167290072-167290073, 167290092, 167290098, 167290114, 167290117-167290118, 167290120, 167290129-167290130, 167290151, 167290223-167290224, 167290228, 167290231, 167290243, 167290262, 167290267, 167290276, 167290288, 167290295, 167290306, 167290332, 167290351, 167290391, 167290419, 167290423, 167290426, 170890378, 170890380, 1723754, 1730045, 1730635, 1730713, 1730752, 1730804, 1730808, 1730821, 1730846-1730849, 1730880, 1730896, 1790149, 1790195, 1790229, 181110005, 181110007, 181110011, 181110013, 1860524, 1860527-1860528, 1860852, 1860855, 1860857, 1875526, 205360322, 2200522, 22980, 231550179, 231550213-231550214, 231550261, 231550483-231550484, 231550528-231550529, 231550649, 231550685-231550689, 23977, 240590-240591, 240596-240597, 240654, 240656, 242010101, 242010237, 243380050, 24483, 245820, 245824, 245840-245841, 245860, 245917, 24815, 250210202-250210209, 250210211-250210215, 250210221-250210222, 250210230-250210231, 250210233-250210237, 250210239, 250210241-250210242, 250210245-250210246, 250210451-250210455, 250210462, 250210824, 25337, 26216, 27501-27502, 27623, 27640, 27669, 27678, 27716, 302378900, 30524, 30527-30528, 30852, 30855, 30857, 3100482, 3100512, 3100657, 3100668, 3100679, 3100720, 3101349-3101350, 3104500, 3104611, 3104700, 3107720, 3107810, 3107820, 3107830, 3107840, 32291, 32327-32328, 33734, 33772, 33774, 3380063, 3380067, 3380080, 3380086, 3383991, 3383993, 34522, 3782245-3782246, 3782511-3782512, 3783096-3783098, 3783266, 3785260-3785265, 3786920, 3786924, 3786955, 3787131-3787133, 398222100, 398222120, 398222180, 398222200, 400330202, 400330215, 400330223, 400330231, 400510604-400510609, 4090124, 4090181-4090183, 4090185-4090187, 4090201, 4090302, 4090323, 4090332, 4090366-4090369, 4090801, 4091112, 41100-41101, 416160150-416160151, 416160176, 416160178, 416160284, 416160300, 420230149, 422360001-422360002, 422380111, 422910024, 422910166-422910167, 422910190-422910191, 422910351-422910352, 422920007, 422920043-422920044, 422920051-422920053, 422920057, 423670121, 423670520-423670521, 423880011-423880013, 423880023-423880025, 424270002, 426050015, 426050025-426050026, 426050031-426050033, 426050044, 426080009, 426580007, 426580010, 426580019, 426580021, 427370101-427370106, 427370110, 427470327, 427910100-427910101, 430660001, 430660006, 430660010, 430660014, 430660018, 430660134, 430660138, 430660201-430660202, 430660252-430660254, 430660301, 430660502, 430660601, 430660610, 430660811, 435980258-435980259, 435980262, 435980283, 435980305, 435980309, 435980344-435980345, 435980348, 435980358, 435980389, 435980392, 435980427, 435980465, 435980541, 435980610-435980611, 435980650, 435980678, 435980682-435980683, 436240001-436240002, 438170906, 439750252-439750257, 439750307-439750308, 439750315, 440873535, 440874000, 440876000, 444953, 445670504-445670507, 445670509- |

| | |
|---|---|
| | 445670511, 445670530, 459630607-459630609, 459630611-459630615, 459630619-459630621, 459630623-459630624, 459630636-459630638, 459630640, 459630686, 459630733-459630734, 459630765, 459630781, 459630790, 460140296, 460141195, 460260983, 4690125, 4690625, 4690725, 4691425, 473350046-473350047, 473350049-473350050, 473350082-473350083, 473350150-473350151, 473350153-473350154, 473350176, 473350178, 473350284-473350286, 473350300, 473350303, 473350323, 473350361, 473350401, 473350472, 473350475, 473350890-473350893, 473350895, 473350929-473350930, 473350937, 473350939, 473350953, 473510001, 473510004, 473510006-473510007, 473510009, 473510017, 473510024-473510027, 473510029, 473510032-473510033, 473510036-473510038, 473510057, 473510075, 473510080, 477810200, 477810256, 477810591-477810595, 477810603-477810606, 477810609-477810610, 488180001, 493150003, 493150005, 493150007-493150009, 498840119, 498840125, 498840127, 501110965-501110967, 502420051, 502420053, 502420060-502420064, 502420070, 502420077, 502420087-502420088, 502420090-502420091, 502420094, 502420103, 502420105, 502420108-502420109, 502420130, 502420132, 502420134, 502420140, 502420145, 502420245, 502420260, 502420333, 502420717, 502420917-502420918, 502680154, 502680426-502680427, 502680761-502680763, 504190171, 504190357, 504190385, 504190390-504190392, 504190395, 504190488, 507420401-507420402, 507420404-507420406, 507420420, 507420423, 507420427-507420428, 507420430-507420431, 507420438, 507420445-507420448, 507420463, 507420477, 507420481-507420483, 507420496-507420498, 507420512, 507420519-507420520, 508810005, 508810010, 508810015, 508810020, 508810025-508810028, 510230303-510230305, 510230319, 510230327, 510230363, 510230366-510230368, 510230801, 510790510, 511440001-511440002, 511440020, 511440030, 511440050, 514070095-514070096, 514070181, 514070269-514070270, 5170920, 5170950, 5171910, 5171920, 5171940, 518170058, 518170170, 518620083-518620088, 519910218-519910219, 519910376-519910377, 519910797, 519910890-519910892, 519910922-519910923, 519910936-519910938, 522660014, 526090001, 526093001, 5271777-5271782, 528180001-528180002, 531040151, 531040211, 531500247, 531500250, 531500314-531500315, 531500317, 531500320, 531500336, 531500386, 531500411, 533990811, 538080411, 538330011-538330016, 538690187-538690189, 538690262, 538692230-538692231, 538692323, 539640001-539640002, 540248-540249, 540271-540272, 540320-540325, 540382-540383, 540480-540481, 540497, 540870305, 540870319, 540870327, 540870339, 540870342, 540870363, 542880106, 544129-544130, 544820053-544820054, 544820301, 548681126, 548684142-548684143, 548684339, 548685005, 548685218, 548685260, 548685289-548685290, 548685348, 548685350, 548685354, 548685427, 548685447, 548685474, 548685596, 548685759, 548685980, 548790014, 548790021-548790022, 548790027, 548790036, 549730605, 549732905, 551110496-551110497, 551110556, 551110686-551110687, 551350132, 551500331-551500335, 551500352-551500354, 551500386, 552920811, 552920911, 553900005-553900006, 553900030, 553900047-553900048, 553900069-553900070, 553900083-553900085, 553900090-553900091, 553900099, 553900108, 553900112, 553900114-553900115, 553900124, 553900131-553900135, 553900142, 553900150-553900156, 553900187, 553900207-553900208, 553900231-553900233, 553900235-553900238, 553900241-553900248, 553900281, 553900291-553900293, 553900304, 553900314, 553900337, 553900339, 553900370, 553900391, 553900414, 553900491-553900493, 553900805-553900809, 555130078-555130079, 555130132, 555130141, 555130160, 555130206-555130207, 555130224, 555130326, 555130954-555130956, 556480632-556480636, 572770001-572770002, 572770105-572770107, 576650331, 578843001-578843003, 578843021, 578843041-578843043, 578843051-578843052, 578843071-578843073, 578843121, 578844001-578844002, 578940150, 578940155, 578940184, 578940195, 578940199, 578940420-578940421, 578940502-578940503, 579020249, 579620014, 579620070, 579620140, 579620280, 579620420, 579620560, 580630001, 581813030-581813032, 581813040-581813043, 582430031, 584680080, 584680100, 584680200, 584680357, 584687820, 584687840, 5902291, 5902327-5902328, 5903772, 5903774, 5904522, 5912897, 5913562-5913563, 591480046-591480047, 591480070, 5915019, 595720102, 595720205, 595720210, 595720215, 595720220, 595720302, 595720402, 595720405, 595720410, 595720415, 595720420, 595720425, 595720501-595720504, 595720705, 595720710, 595720720, 595720730, 595720740, 595720983-595720984, 596510204-596510205, 596510240-596510241, 596760030, 596760040, 596760050, 596760201, 596760600, 596760610, 596760960, 596760966, 5970137-5970138, 5970141, 597625091, 597625093, 597650048-597650051, 597650072-597650073, 597650098, 597650129-597650130, 597650224, 597650231, 597650267, 597650276, 597650288, 597650295, 597650306, 597650332, 599230604, 599230701-599230716, 599230721-599230727, 604290925-604290926, 605052900-605052901, 605053628-605053631, 605054327, 605056113-605056115, 605056128, 605056132, 605056166, 605056177, 60568, 606870149, 606870192, 606870203, 606870455, 611260003-611260004, 611260103, 611260507, 611260509-611260511, 611260514-611260517, 612690410, 617030303-617030305, 617030309, 617030319, 617030323, 617030327, 617030331-617030332, 617030339, 617030341-617030344, 617030347-617030349, 617030359-617030363, 617550007-617550008, 619581701-619581702, 621064878, 621750240-621750245, 621950072, 621950200, 621950210, 621950610-621950612, 625590920-625590925, 627560008, 627560023, 627560073, 627560102, 627560219, 627560238-627560239, 627560321, 627560438, 627560452, 627560533, 627560614, 627560746, 627560826-627560827, 627560974, 628560001, 628560177, 628560389, 628560600, 628560602-628560604, 628560704, 628560708, 628560710, 628560712, 628560714, 628560718, 628560720, 628560724, 630200049, 630200078-630200080, 630200090, 630200113, 630200180, 630200198, 630200230, 630200390, 630200400, 630200533-630200535, 63026, 63029, 633040095-633040096, 633040135, 633230101-633230104, 633230117, 633230120, 633230125-633230128, 633230132, 633230136-633230137, 633230140, 633230142, 633230145, 633230151, 633230172, 633230174-633230176, 633230192-633230194, 633230196, 633230211-633230212, 633230278, |

| | |
|---|---|
| | 633230365, 633230376-633230379, 633230572, 633230637, 633230650, 633230715, 633230721, 633230750, 633230760, 633230762-633230763, 633230771, 633230825, 633230883, 633790011, 633790023, 634590103-634590104, 634590177, 634590303, 634590305, 634590348, 634590390-634590391, 634590395-634590396, 634590600-634590601, 635390017, 635390019, 635390026, 635390044, 635390117, 635390187-635390189, 635390197, 635390284, 635390295, 635390486, 635390688, 637390932, 637390971, 637590003, 637593000-637593001, 641160011, 641440501-641440506, 6416174-6416178, 643700210, 643700250, 643700532, 646790021, 646790067, 646790096, 646790632-646790636, 646790793-646790794, 648420727, 648421020, 648421025, 649800276-649800277, 649800333-649800338, 649800404, 649800418, 65033, 650447462-650447463, 651620794-651620795, 651620801-651620806, 651620843-651620844, 652190160, 652190200, 653925000, 654830116, 655970402, 655970406, 662150016, 663020014, 664350410, 667330948, 667330958, 667580042-667580048, 667580050-667580051, 667580053-667580055, 667580950, 668280030, 669142327-669142328, 669930489, 671840501-671840502, 671840508-671840513, 673860411, 673860811, 674570195, 674570215, 674570238-674570239, 674570245-674570246, 674570254, 674570268, 674570311, 674570316, 674570325-674570326, 674570357-674570358, 674570393-674570396, 674570424-674570425, 674570429, 674570431, 674570434, 674570436, 674570442-674570443, 674570449-674570452, 674570454-674570455, 674570462-674570464, 674570468-674570469, 674570471, 674570474, 674570476, 674570478-674570479, 674570481-674570482, 674570491-674570495, 674570513, 674570531-674570533, 674570546, 674570579, 674570608-674570609, 674570615-674570618, 674570662, 674570781, 674570845, 674570847, 674570893, 674570928, 674570991, 676510323, 678770458-678770459, 678770537-678770542, 678770633-678770634, 679790001, 680010265-680010266, 680010282-680010284, 680010313, 680010341-680010342, 680010345, 680010347-680010348, 680010350, 680010359, 680010370-680010372, 680010422, 680010424-680010426, 680010442-680010444, 680010468, 680010480, 680830148-680830149, 680830162-680830163, 680830170-680830171, 680830176-680830180, 680830190-680830193, 680830202, 680830248-680830250, 680830259, 680830269-680830272, 680830292-680830293, 680830314, 680830326-680830328, 680830337, 680830343-680830346, 680830361, 680830381-680830382, 680830386, 681520103, 681520106, 681520108-681520109, 681800390-681800391, 681800738, 681800801, 683820751-683820756, 683820913-683820915, 683820997, 684280033, 684280083, 684280225, 684620317, 684750503, 686250811, 686820003, 687270712, 687270745, 688170134, 688420301, 690067, 690070, 690074-690076, 690078-690081, 690084, 690086, 690099, 690103, 690135-690136, 690145, 690151-690155, 690169-690171, 690173-690174, 690176, 690187-690189, 690193, 690197, 690201, 690205, 690227, 690231, 690238, 690249, 690284, 690296, 690298, 690305, 690315, 690342, 690486, 690550, 690688, 690770, 690830, 690970239-690970243, 690970262, 690970274, 690970285, 690970313-690970314, 690970346, 690970353, 690970359, 690970364, 690970368-690970369, 690970371-690970372, 690970501, 690970516-690970517, 690970572, 690970594, 690970635-690970636, 690970805, 690970948-690970949, 690980, 690983, 691010, 691195, 691198, 691531, 691710398, 691890063, 691890382-691890383, 691890403, 691897638, 692114000, 692299, 692381165, 692381250, 693030-693034, 693857-693859, 694004, 694015, 694026, 694030-694034, 694037, 694480005, 694495-694496, 694880003, 695390019-695390020, 695390049, 695390115, 695390123-695390124, 695390152, 695390157-695390159, 696560103, 696600201-696600203, 698140-698141, 698800120, 699141-699142, 699144, 699180720, 699321, 700201910-700201911, 701211218-701211219, 701211221-701211223, 701211236, 701211238-701211240, 701211244, 701211463, 701211482, 701211567, 701211630-701211631, 701211644, 702550010, 702550020, 702550025, 702970003-702970004, 7033015, 7033018-7033019, 7033067, 7033069, 7033071, 7033125, 7033154-7033155, 7033213, 7033216-7033218, 7033249, 7033301, 7033311, 7033321, 7033333, 7033343, 7033427, 7033429, 7033985-7033986, 7034004, 7034100, 7034106, 7034116, 7034154-7034156, 7034182-7034183, 7034239, 7034244, 7034246, 7034248, 7034402, 7034412, 7034432, 7034434, 7034636, 7034680, 7034685-7034686, 7034714, 7034764, 7034766-7034768, 7034852, 7035040, 7035043, 7035046, 7035075, 7035233, 7035653, 7035656-7035657, 7035720, 7035730, 7035747-7035748, 7035775, 7035778, 7035854, 704370240, 705340002, 707000169-707000170, 707101525, 707101530-707101531, 707101610, 707560815-707560816, 707711092-707711097, 707711521-707711523, 708350003-708350004, 708600200-708600201, 708600204-708600206, 708600208, 708600211, 708600214-708600218, 712580015, 712870119, 712870219, 712880100-712880101, 712880108-712880109, 712880112-712880116, 712880127-712880129, 712880555, 713340100, 717316121, 717770390-717770392, 717790115, 717790125, 719730090, 720640110, 720640120, 720640130, 720640210, 721870401, 722050006-722050007, 722050030-722050031, 722050036, 722050045-722050046, 722050050, 722370101, 722660108, 722660125-722660126, 722660128, 722660161-722660162, 724850201-724850205, 724850210-724850219, 724850221-724850223, 725790011, 726030101, 726030103-726030105, 726030107, 726030200, 726030301, 726030326, 726030411, 726060554-726060559, 726060566, 726070100, 726110779, 726110785, 726940515, 726940954, 728930001-728930003, 728930005, 728930007-728930008, 730900420-730900421, 731160100-731160102, 732070101, 73260-73262, 733804700, 735350208, 740561, 740566, 740576, 740579, 74201, 74205, 74207, 74401, 758001, 758003-758005, 759870111, 760750101-760750103, 761350005-761350006, 761350009-761350011, 761890113, 761890533-761890535, 763100022, 763360080, 763880001, 763880635, 763880713, 763880880, 764721132, 780180-780184, 780401, 780438, 780495, 780526, 780566-780567, 780592, 780594, 780620, 780626-780628, 780640, 780645-780652, 780666, 780668-780674, 780681-780683, 780690, 780694, 780698, 780701, 780708-780709, 780715-780716, 780811, 780818, 780825, 780846, 780860, 780867, 780874, 780909, 780916, 780923, 780951, 780958, 781007, 7812691-7812696, 7813079, 7813139, 7813164-7813168, 7813233, 7813244, 7813253, 7813255, 7813282-7813283, 7813296, 7813315, 7813317, 7813492, |

| | |
|---|---|
| | 7819253, 7819315, 7819317, 796720018-796720019, 796720825-796720826, 80045, 80100, 81179, 8320285, 84510, 851366, 851381, 851417, 851425, 851430, 851519, 853004, 9046621, 9046901, 9046948, 91111, 931125, 937473-937474, 937599-937602, 937629-937630, 937638-937639, 937663-937664, 937766-937768, 9443810, 9443922, 95091, 95093, 9551020-9551022, 9551725, 9551727, 9551731, 9551733, 9551746, 97224, 97529 |
| Radiation | **HCPCS:** 63620-63621, 67218, 77014, 77371-77373, 77385-77387, 77401-77404, 77406-77409, 77411-77414, 77416, 77418, 77421-77425, 77469, 77520, 77522-77523, 77525, 77750, 77761-77763, 77767-77768, 77770-77772, 77776-77778, 77785-77787, 77789, 79200, 79300, 79403, 79440, 79445, 0394T, 0395T, G6002, G6003, G6004, G6005, G6006, G6007, G6008, G6009, G6010, G6011, G6012, G6013, G6014, G6015, G6016, G6017, G9894, Q3001<br><br>**ICD-9-PCS:** 142.6, 922.0-923.3, 923.9, 924.1<br><br>**ICD-10-PCS:** 085E3ZZ, 085F3ZZ, 08H031Z, 08H0X1Z, 08H131Z, 08H1X1Z, 0BH001Z, 0BH031Z, 0BH041Z, 0BH071Z, 0BH081Z, 0BHK01Z, 0BHK31Z, 0BHK41Z, 0BHK71Z, 0BHK81Z, 0BHL01Z, 0BHL31Z, 0BHL41Z, 0BHL71Z, 0BHL81Z, 0CH701Z, 0CH731Z, 0CH7X1Z, 0DH501Z, 0DH531Z, 0DH541Z, 0DH571Z, 0DH581Z, 0DHP01Z, 0DHP31Z, 0DHP41Z, 0DHP71Z, 0DHP81Z, 0FHB01Z, 0FHB31Z, 0FHB41Z, 0FHB71Z, 0FHB81Z, 0FHD01Z, 0FHD31Z, 0FHD41Z, 0FHD71Z, 0FHD81Z, 0HHT01Z, 0HHT31Z, 0HHT71Z, 0HHT81Z, 0HHTX1Z, 0HHU01Z, 0HHU31Z, 0HHU71Z, 0HHU81Z, 0HHUX1Z, 0HHV01Z, 0HHV31Z, 0HHV71Z, 0HHV81Z, 0HHVX1Z, 0HHW01Z, 0HHW31Z, 0HHW71Z, 0HHW81Z, 0HHWX1Z, 0HHX01Z, 0HHX31Z, 0HHX71Z, 0HHX81Z, 0HHXX1Z, 0JHS01Z, 0JHS31Z, 0JHT01Z, 0JHT31Z, 0JHV01Z, 0JHV31Z, 0JHW01Z, 0JHW31Z, 0UHC01Z, 0UHC31Z, 0UHC41Z, 0UHC71Z, 0UHC81Z, 0UHG01Z, 0UHG31Z, 0UHG41Z, 0UHG71Z, 0UHG81Z, 0UHGX1Z, 0VH001Z, 0VH031Z, 0VH041Z, 0VH071Z, 0VH081Z, 0WH001Z, 0WH031Z, 0WH041Z, 0WH101Z, 0WH131Z, 0WH141Z, 0WH201Z, 0WH231Z, 0WH241Z, 0WH301Z, 0WH331Z, 0WH341Z, 0WH401Z, 0WH431Z, 0WH441Z, 0WH501Z, 0WH531Z, 0WH541Z, 0WH601Z, 0WH631Z, 0WH641Z, 0WH801Z, 0WH831Z, 0WH841Z, 0WH901Z, 0WH931Z, 0WH941Z, 0WHB01Z, 0WHB31Z, 0WHB41Z, 0WHC01Z, 0WHC31Z, 0WHC41Z, 0WHD01Z, 0WHD31Z, 0WHD41Z, 0WHF01Z, 0WHF31Z, 0WHF41Z, 0WHG01Z, 0WHG31Z, 0WHG41Z, 0WHH01Z, 0WHH31Z, 0WHH41Z, 0WHJ01Z, 0WHJ31Z, 0WHJ41Z, 0WHK01Z, 0WHK31Z, 0WHK41Z, 0WHL01Z, 0WHL31Z, 0WHL41Z, 0WHM01Z, 0WHM31Z, 0WHM41Z, 0WHN01Z, 0WHN31Z, 0WHN41Z, 0WHP01Z, 0WHP31Z, 0WHP41Z, 0WHP71Z, 0WHP81Z, 0WHQ01Z, 0WHQ31Z, 0WHQ41Z, 0WHQ71Z, 0WHQ81Z, 0WHR01Z, 0WHR31Z, 0WHR41Z, 0WHR71Z, 0WHR81Z, 0XH201Z, 0XH231Z, 0XH241Z, 0XH301Z, 0XH331Z, 0XH341Z, 0XH401Z, 0XH431Z, 0XH441Z, 0XH501Z, 0XH531Z, 0XH541Z, 0XH601Z, 0XH631Z, 0XH641Z, 0XH701Z, 0XH731Z, 0XH741Z, 0XH801Z, 0XH831Z, 0XH841Z, 0XH901Z, 0XH931Z, 0XH941Z, 0XHB01Z, 0XHB31Z, 0XHB41Z, 0XHC01Z, 0XHC31Z, 0XHC41Z, 0XHD01Z, 0XHD31Z, 0XHD41Z, 0XHF01Z, 0XHF31Z, 0XHF41Z, 0XHG01Z, 0XHG31Z, 0XHG41Z, 0XHH01Z, 0XHH31Z, 0XHH41Z, 0XHJ01Z, 0XHJ31Z, 0XHJ41Z, 0XHK01Z, 0XHK31Z, 0XHK41Z, 0YH001Z, 0YH031Z, 0YH041Z, 0YH101Z, 0YH131Z, 0YH141Z, 0YH501Z, 0YH531Z, 0YH541Z, 0YH601Z, 0YH631Z, 0YH641Z, 0YH701Z, 0YH731Z, 0YH741Z, 0YH801Z, 0YH831Z, 0YH841Z, 0YH901Z, 0YH931Z, 0YH941Z, 0YHB01Z, 0YHB31Z, 0YHB41Z, 0YHC01Z, 0YHC31Z, 0YHC41Z, 0YHD01Z, 0YHD31Z, 0YHD41Z, 0YHF01Z, 0YHF31Z, 0YHF41Z, 0YHG01Z, 0YHG31Z, 0YHG41Z, 0YHH01Z, 0YHH31Z, 0YHH41Z, 0YHJ01Z, 0YHJ31Z, 0YHJ41Z, 0YHK01Z, 0YHK31Z, 0YHK41Z, 0YHL01Z, 0YHL31Z, 0YHL41Z, 0YHM01Z, 0YHM31Z, 0YHM41Z, 0YHN01Z, 0YHN31Z, 0YHN41Z, 3E0B304, 3E0B704, 3E0BX04, 3E0C304, 3E0C704, 3E0CX04, 3E0D304, 3E0D704, 3E0DX04, 3E0E304, 3E0E704, 3E0E804, 3E0F304, 3E0F704, 3E0F804, 3E0G304, 3E0G704, 3E0G804, 3E0H304, 3E0H704, 3E0H804, 3E0J304, 3E0J704, 3E0J804, 3E0K304, 3E0K704, 3E0K804, 3E0L304, 3E0L704, 3E0M304, 3E0M704, 3E0N304, 3E0N704, 3E0N804, 3E0P304, 3E0P704, 3E0P804, 3E0Q004, 3E0Q0HZ, 3E0Q304, 3E0Q704, 3E0R304, 3E0S304, 3E0U304, 3E0Y304, 3E0Y704, CW70NZZ, CW70YZZ, CW73NZZ, CW73YZZ, CW7GGZZ, CW7GYZZ, CW7N8ZZ, CW7NGZZ, CW7NNZZ, CW7NPZZ, CW7NYZZ, CW7YYZZ, D0000ZZ, D0001ZZ, D0002ZZ, D0003Z0, D0003ZZ, D0004ZZ, D0005ZZ, D0010ZZ, D0011ZZ, D0012ZZ, D0013Z0, D0013ZZ, D0014ZZ, D0015ZZ, D0060ZZ, D0061ZZ, D0062ZZ, D0063Z0, D0063ZZ, D0064ZZ, D0065ZZ, D0070ZZ, D0071ZZ, D0072ZZ, D0073Z0, D0073ZZ, D0074ZZ, D0075ZZ, D01097Z, D01098Z, D01099Z, D0109BZ, D0109CZ, D0109YZ, D010B7Z, D010B8Z, D010B9Z, D010BB1, D010BBZ, D010BCZ, D010BYZ, D01197Z, D01198Z, D01199Z, D0119BZ, D0119CZ, D0119YZ, D011B7Z, D011B8Z, D011B9Z, D011BB1, D011BBZ, D011BCZ, D011BYZ, D01697Z, D01698Z, D01699Z, D0169BZ, D0169CZ, D0169YZ, D016B7Z, D016B8Z, D016B9Z, D016BB1, D016BBZ, D016BCZ, D016BYZ, D01797Z, D01798Z, D01799Z, D0179BZ, D0179CZ, D0179YZ, D017B7Z, D017B8Z, D017B9Z, D017BB1, D017BBZ, D017BCZ, D017BYZ, D020DZZ, D020HZZ, D020JZZ, D021DZZ, D021HZZ, D021JZZ, D027DZZ, D027HZZ, D027JZZ, D0Y07ZZ, D0Y0FZZ, D0Y17ZZ, D0Y1FZZ, D0Y67ZZ, D0Y6FZZ, D0Y77ZZ, D0Y7FZZ, D7000ZZ, D7001ZZ, D7002ZZ, D7003Z0, D7003ZZ, D7004ZZ, D7005ZZ, D7010ZZ, D7011ZZ, D7012ZZ, D7013Z0, D7013ZZ, D7014ZZ, D7015ZZ, D7020ZZ, D7021ZZ, D7022ZZ, D7023Z0, D7023ZZ, D7024ZZ, D7025ZZ, D7030ZZ, D7031ZZ, D7032ZZ, D7033Z0, D7033ZZ, D7034ZZ, D7035ZZ, D7040ZZ, D7041ZZ, D7042ZZ, D7043Z0, D7043ZZ, D7044ZZ, D7045ZZ, D7050ZZ, D7051ZZ, D7052ZZ, D7053Z0, D7053ZZ, D7054ZZ, D7055ZZ, D7060ZZ, D7061ZZ, D7062ZZ, D7063Z0, D7063ZZ, D7064ZZ, D7065ZZ, D7070ZZ, D7071ZZ, D7072ZZ, D7073Z0, D7073ZZ, D7074ZZ, D7075ZZ, D7080ZZ, D7081ZZ, D7082ZZ, D7083Z0, D7083ZZ, D7084ZZ, D7085ZZ, D71097Z, D71098Z, D71099Z, D7109BZ, D7109CZ, D7109YZ, D710B7Z, D710B8Z, D710B9Z, D710BB1, D710BBZ, D710BCZ, D710BYZ, D71197Z, D71198Z, D71199Z, D7119BZ, D7119CZ, D7119YZ, D711B7Z, D711B8Z, D711B9Z, D711BB1, D711BBZ, D711BCZ, D711BYZ, D71297Z, D71298Z, |

| | |
|---|---|
| | D71299Z, D7129BZ, D7129CZ, D7129YZ, D712B7Z, D712B8Z, D712B9Z, D712BB1, D712BBZ, D712BCZ, D712BYZ, D71397Z, D71398Z, D71399Z, D7139BZ, D7139CZ, D7139YZ, D713B7Z, D713B8Z, D713B9Z, D713BB1, D713BBZ, D713BCZ, D713BYZ, D71497Z, D71498Z, D71499Z, D7149BZ, D7149CZ, D7149YZ, D714B7Z, D714B8Z, D714B9Z, D714BB1, D714BBZ, D714BCZ, D714BYZ, D71597Z, D71598Z, D71599Z, D7159BZ, D7159CZ, D7159YZ, D715B7Z, D715B8Z, D715B9Z, D715BB1, D715BBZ, D715BCZ, D715BYZ, D71697Z, D71698Z, D71699Z, D7169BZ, D7169CZ, D7169YZ, D716B7Z, D716B8Z, D716B9Z, D716BB1, D716BBZ, D716BCZ, D716BYZ, D71797Z, D71798Z, D71799Z, D7179BZ, D7179CZ, D7179YZ, D717B7Z, D717B8Z, D717B9Z, D717BB1, D717BBZ, D717BCZ, D717BYZ, D71897Z, D71898Z, D71899Z, D7189BZ, D7189CZ, D7189YZ, D718B7Z, D718B8Z, D718B9Z, D718BB1, D718BBZ, D718BCZ, D718BYZ, D7Y0FZZ, D7Y1FZZ, D7Y2FZZ, D7Y3FZZ, D7Y4FZZ, D7Y5FZZ, D7Y6FZZ, D7Y7FZZ, D7Y8FZZ, D8000ZZ, D8001ZZ, D8002ZZ, D8003Z0, D8003ZZ, D8004ZZ, D8005ZZ, D81097Z, D81098Z, D81099Z, D8109BZ, D8109CZ, D8109YZ, D810B7Z, D810B8Z, D810B9Z, D810BB1, D810BBZ, D810BCZ, D810BYZ, D8Y07ZZ, D8Y0FZZ, D9000ZZ, D9001ZZ, D9002ZZ, D9003Z0, D9003ZZ, D9004ZZ, D9005ZZ, D9010ZZ, D9011ZZ, D9012ZZ, D9013Z0, D9013ZZ, D9014ZZ, D9015ZZ, D9030ZZ, D9031ZZ, D9032ZZ, D9033Z0, D9033ZZ, D9034ZZ, D9035ZZ, D9040ZZ, D9041ZZ, D9042ZZ, D9043Z0, D9043ZZ, D9044ZZ, D9045ZZ, D9050ZZ, D9051ZZ, D9052ZZ, D9053Z0, D9053ZZ, D9054ZZ, D9055ZZ, D9060ZZ, D9061ZZ, D9062ZZ, D9063Z0, D9063ZZ, D9064ZZ, D9065ZZ, D9070ZZ, D9071ZZ, D9072ZZ, D9073Z0, D9073ZZ, D9074ZZ, D9075ZZ, D9080ZZ, D9081ZZ, D9082ZZ, D9083Z0, D9083ZZ, D9084ZZ, D9085ZZ, D9090ZZ, D9091ZZ, D9092ZZ, D9093Z0, D9093ZZ, D9094ZZ, D9095ZZ, D90B0ZZ, D90B1ZZ, D90B2ZZ, D90B3Z0, D90B3ZZ, D90B4ZZ, D90B5ZZ, D90D0ZZ, D90D1ZZ, D90D2ZZ, D90D3Z0, D90D3ZZ, D90D4ZZ, D90D5ZZ, D90F0ZZ, D90F1ZZ, D90F2ZZ, D90F3Z0, D90F3ZZ, D90F4ZZ, D90F5ZZ, D91097Z, D91098Z, D91099Z, D9109BZ, D9109CZ, D9109YZ, D910B7Z, D910B8Z, D910B9Z, D910BB1, D910BBZ, D910BCZ, D910BYZ, D91197Z, D91198Z, D91199Z, D9119BZ, D9119CZ, D9119YZ, D911B7Z, D911B8Z, D911B9Z, D911BB1, D911BBZ, D911BCZ, D911BYZ, D91397Z, D91398Z, D91399Z, D9139BZ, D9139CZ, D9139YZ, D913B7Z, D913B8Z, D913B9Z, D913BB1, D913BBZ, D913BCZ, D913BYZ, D91497Z, D91498Z, D91499Z, D9149BZ, D9149CZ, D9149YZ, D914B7Z, D914B8Z, D914B9Z, D914BB1, D914BBZ, D914BCZ, D914BYZ, D91597Z, D91598Z, D91599Z, D9159BZ, D9159CZ, D9159YZ, D915B7Z, D915B8Z, D915B9Z, D915BB1, D915BBZ, D915BCZ, D915BYZ, D91697Z, D91698Z, D91699Z, D9169BZ, D9169CZ, D9169YZ, D916B7Z, D916B8Z, D916B9Z, D916BB1, D916BBZ, D916BCZ, D916BYZ, D91797Z, D91798Z, D91799Z, D9179BZ, D9179CZ, D9179YZ, D917B7Z, D917B8Z, D917B9Z, D917BB1, D917BBZ, D917BCZ, D917BYZ, D91897Z, D91898Z, D91899Z, D9189BZ, D9189CZ, D9189YZ, D918B7Z, D918B8Z, D918B9Z, D918BB1, D918BBZ, D918BCZ, D918BYZ, D91997Z, D91998Z, D91999Z, D9199BZ, D9199CZ, D9199YZ, D919B7Z, D919B8Z, D919B9Z, D919BB1, D919BBZ, D919BCZ, D919BYZ, D91B97Z, D91B98Z, D91B99Z, D91B9BZ, D91B9CZ, D91B9YZ, D91BB7Z, D91BB8Z, D91BB9Z, D91BBB1, D91BBBZ, D91BBCZ, D91BBYZ, D91D97Z, D91D98Z, D91D99Z, D91D9BZ, D91D9CZ, D91D9YZ, D91DB7Z, D91DB8Z, D91DB9Z, D91DBB1, D91DBBZ, D91DBCZ, D91DBYZ, D91F97Z, D91F98Z, D91F99Z, D91F9BZ, D91F9CZ, D91F9YZ, D91FB7Z, D91FB8Z, D91FB9Z, D91FBB1, D91FBBZ, D91FBCZ, D91FBYZ, D9Y07ZZ, D9Y0FZZ, D9Y17ZZ, D9Y1FZZ, D9Y37ZZ, D9Y47ZZ, D9Y4CZZ, D9Y4FZZ, D9Y57ZZ, D9Y5FZZ, D9Y67ZZ, D9Y6FZZ, D9Y77ZZ, D9Y7FZZ, D9Y87ZZ, D9Y8FZZ, D9Y97ZZ, D9Y9FZZ, D9YB7ZZ, D9YBCZZ, D9YBFZZ, D9YCCZZ, D9YCFZZ, D9YD7ZZ, D9YDCZZ, D9YDFZZ, D9YF7ZZ, DB000ZZ, DB001ZZ, DB002ZZ, DB003Z0, DB003ZZ, DB004ZZ, DB005ZZ, DB010ZZ, DB011ZZ, DB012ZZ, DB013Z0, DB013ZZ, DB014ZZ, DB015ZZ, DB020ZZ, DB021ZZ, DB022ZZ, DB023Z0, DB023ZZ, DB024ZZ, DB025ZZ, DB050ZZ, DB051ZZ, DB052ZZ, DB053Z0, DB053ZZ, DB054ZZ, DB055ZZ, DB060ZZ, DB061ZZ, DB062ZZ, DB063Z0, DB063ZZ, DB064ZZ, DB065ZZ, DB070ZZ, DB071ZZ, DB072ZZ, DB073Z0, DB073ZZ, DB074ZZ, DB075ZZ, DB080ZZ, DB081ZZ, DB082ZZ, DB083Z0, DB083ZZ, DB084ZZ, DB085ZZ, DB1097Z, DB1098Z, DB1099Z, DB109BZ, DB109CZ, DB109YZ, DB10B7Z, DB10B8Z, DB10B9Z, DB10BB1, DB10BBZ, DB10BCZ, DB10BYZ, DB1197Z, DB1198Z, DB1199Z, DB119BZ, DB119CZ, DB119YZ, DB11B7Z, DB11B8Z, DB11B9Z, DB11BB1, DB11BBZ, DB11BCZ, DB11BYZ, DB1297Z, DB1298Z, DB1299Z, DB129BZ, DB129CZ, DB129YZ, DB12B7Z, DB12B8Z, DB12B9Z, DB12BB1, DB12BBZ, DB12BCZ, DB12BYZ, DB1597Z, DB1598Z, DB1599Z, DB159BZ, DB159CZ, DB159YZ, DB15B7Z, DB15B8Z, DB15B9Z, DB15BB1, DB15BBZ, DB15BCZ, DB15BYZ, DB1697Z, DB1698Z, DB1699Z, DB169BZ, DB169CZ, DB169YZ, DB16B7Z, DB16B8Z, DB16B9Z, DB16BB1, DB16BBZ, DB16BCZ, DB16BYZ, DB1797Z, DB1798Z, DB1799Z, DB179BZ, DB179CZ, DB179YZ, DB17B7Z, DB17B8Z, DB17B9Z, DB17BB1, DB17BBZ, DB17BCZ, DB17BYZ, DB1897Z, DB1898Z, DB1899Z, DB189BZ, DB189CZ, DB189YZ, DB18B7Z, DB18B8Z, DB18B9Z, DB18BB1, DB18BBZ, DB18BCZ, DB18BYZ, DBY07ZZ, DBY0FZZ, DBY17ZZ, DBY1FZZ, DBY27ZZ, DBY2FZZ, DBY57ZZ, DBY5FZZ, DBY67ZZ, DBY6FZZ, DBY77ZZ, DBY7FZZ, DBY87ZZ, DBY8FZZ, DD000ZZ, DD001ZZ, DD002ZZ, DD003Z0, DD003ZZ, DD004ZZ, DD005ZZ, DD010ZZ, DD011ZZ, DD012ZZ, DD013Z0, DD013ZZ, DD014ZZ, DD015ZZ, DD020ZZ, DD021ZZ, DD022ZZ, DD023Z0, DD023ZZ, DD024ZZ, DD025ZZ, DD030ZZ, DD031ZZ, DD032ZZ, DD033Z0, DD033ZZ, DD034ZZ, DD035ZZ, DD040ZZ, DD041ZZ, DD042ZZ, DD043Z0, DD043ZZ, DD044ZZ, DD045ZZ, DD050ZZ, DD051ZZ, DD052ZZ, DD053Z0, DD053ZZ, DD054ZZ, DD055ZZ, DD070ZZ, DD071ZZ, DD072ZZ, DD073Z0, DD073ZZ, DD074ZZ, DD075ZZ, DD1097Z, DD1098Z, DD1099Z, DD109BZ, DD109CZ, DD109YZ, DD10B7Z, DD10B8Z, DD10B9Z, DD10BB1, DD10BBZ, DD10BCZ, DD10BYZ, DD1197Z, DD1198Z, DD1199Z, DD119BZ, DD119CZ, DD119YZ, DD11B7Z, DD11B8Z, DD11B9Z, DD11BB1, DD11BBZ, DD11BCZ, DD11BYZ, DD1297Z, DD1298Z, DD1299Z, DD129BZ, DD129CZ, DD129YZ, DD12B7Z, DD12B8Z, DD12B9Z, DD12BB1, DD12BBZ, DD12BCZ, DD12BYZ, DD1397Z, DD1398Z, DD1399Z, DD139BZ, DD139CZ, DD139YZ, DD13B7Z, DD13B8Z, DD13B9Z, DD13BB1, DD13BBZ, DD13BCZ, DD13BYZ, DD1497Z, DD1498Z, DD1499Z, DD149BZ, DD149CZ, DD149YZ, DD14B7Z, DD14B8Z, DD14B9Z, |

| | |
|---|---|
| | DD14BB1, DD14BBZ, DD14BCZ, DD14BYZ, DD1597Z, DD1598Z, DD1599Z, DD159BZ, DD159CZ, DD159YZ, DD15B7Z, DD15B8Z, DD15B9Z, DD15BB1, DD15BBZ, DD15BCZ, DD15BYZ, DD1797Z, DD1798Z, DD1799Z, DD179BZ, DD179CZ, DD179YZ, DD17B7Z, DD17B8Z, DD17B9Z, DD17BB1, DD17BBZ, DD17BCZ, DD17BYZ, DDY07ZZ, DDY0FZZ, DDY17ZZ, DDY1CZZ, DDY1FZZ, DDY27ZZ, DDY2CZZ, DDY2FZZ, DDY37ZZ, DDY3CZZ, DDY3FZZ, DDY47ZZ, DDY4CZZ, DDY4FZZ, DDY57ZZ, DDY5CZZ, DDY5FZZ, DDY77ZZ, DDY7CZZ, DDY7FZZ, DDY8CZZ, DDY8FZZ, DF000ZZ, DF001ZZ, DF002ZZ, DF003Z0, DF003ZZ, DF004ZZ, DF005ZZ, DF010ZZ, DF011ZZ, DF012ZZ, DF013Z0, DF013ZZ, DF014ZZ, DF015ZZ, DF020ZZ, DF021ZZ, DF022ZZ, DF023Z0, DF023ZZ, DF024ZZ, DF025ZZ, DF030ZZ, DF031ZZ, DF032ZZ, DF033Z0, DF033ZZ, DF034ZZ, DF035ZZ, DF1097Z, DF1098Z, DF1099Z, DF109BZ, DF109CZ, DF109YZ, DF10B7Z, DF10B8Z, DF10B9Z, DF10BB1, DF10BBZ, DF10BCZ, DF10BYZ, DF1197Z, DF1198Z, DF1199Z, DF119BZ, DF119CZ, DF119YZ, DF11B7Z, DF11B8Z, DF11B9Z, DF11BB1, DF11BBZ, DF11BCZ, DF11BYZ, DF1297Z, DF1298Z, DF1299Z, DF129BZ, DF129CZ, DF129YZ, DF12B7Z, DF12B8Z, DF12B9Z, DF12BB1, DF12BBZ, DF12BCZ, DF12BYZ, DF1397Z, DF1398Z, DF1399Z, DF139BZ, DF139CZ, DF139YZ, DF13B7Z, DF13B8Z, DF13B9Z, DF13BB1, DF13BBZ, DF13BCZ, DF13BYZ, DFY07ZZ, DFY0CZZ, DFY0FZZ, DFY17ZZ, DFY1CZZ, DFY1FZZ, DFY27ZZ, DFY2CZZ, DFY2FZZ, DFY37ZZ, DFY3CZZ, DFY3FZZ, DG000ZZ, DG001ZZ, DG002ZZ, DG003Z0, DG003ZZ, DG005ZZ, DG010ZZ, DG011ZZ, DG012ZZ, DG013Z0, DG013ZZ, DG015ZZ, DG020ZZ, DG021ZZ, DG022ZZ, DG023Z0, DG023ZZ, DG025ZZ, DG040ZZ, DG041ZZ, DG042ZZ, DG043Z0, DG043ZZ, DG045ZZ, DG050ZZ, DG051ZZ, DG052ZZ, DG053Z0, DG053ZZ, DG055ZZ, DG1097Z, DG1098Z, DG1099Z, DG109BZ, DG109CZ, DG109YZ, DG10B7Z, DG10B8Z, DG10B9Z, DG10BB1, DG10BBZ, DG10BCZ, DG10BYZ, DG1197Z, DG1198Z, DG1199Z, DG119BZ, DG119CZ, DG119YZ, DG11B7Z, DG11B8Z, DG11B9Z, DG11BB1, DG11BBZ, DG11BCZ, DG11BYZ, DG1297Z, DG1298Z, DG1299Z, DG129BZ, DG129CZ, DG129YZ, DG12B7Z, DG12B8Z, DG12B9Z, DG12BB1, DG12BBZ, DG12BCZ, DG12BYZ, DG1497Z, DG1498Z, DG1499Z, DG149BZ, DG149CZ, DG149YZ, DG14B7Z, DG14B8Z, DG14B9Z, DG14BB1, DG14BBZ, DG14BCZ, DG14BYZ, DG1597Z, DG1598Z, DG1599Z, DG159BZ, DG159CZ, DG159YZ, DG15B7Z, DG15B8Z, DG15B9Z, DG15BB1, DG15BBZ, DG15BCZ, DG15BYZ, DG20DZZ, DG20HZZ, DG20JZZ, DGY07ZZ, DGY0FZZ, DGY17ZZ, DGY1FZZ, DGY27ZZ, DGY2FZZ, DGY47ZZ, DGY4FZZ, DGY57ZZ, DGY5FZZ, DH020ZZ, DH021ZZ, DH022ZZ, DH023Z0, DH023ZZ, DH024ZZ, DH025ZZ, DH030ZZ, DH031ZZ, DH032ZZ, DH033Z0, DH033ZZ, DH034ZZ, DH035ZZ, DH040ZZ, DH041ZZ, DH042ZZ, DH043Z0, DH043ZZ, DH044ZZ, DH045ZZ, DH060ZZ, DH061ZZ, DH062ZZ, DH063Z0, DH063ZZ, DH064ZZ, DH065ZZ, DH070ZZ, DH071ZZ, DH072ZZ, DH073Z0, DH073ZZ, DH074ZZ, DH075ZZ, DH080ZZ, DH081ZZ, DH082ZZ, DH083Z0, DH083ZZ, DH084ZZ, DH085ZZ, DH090ZZ, DH091ZZ, DH092ZZ, DH093Z0, DH093ZZ, DH094ZZ, DH095ZZ, DH0B0ZZ, DH0B1ZZ, DH0B2ZZ, DH0B3Z0, DH0B3ZZ, DH0B4ZZ, DH0B5ZZ, DHY27ZZ, DHY2FZZ, DHY37ZZ, DHY3FZZ, DHY47ZZ, DHY4FZZ, DHY5FZZ, DHY67ZZ, DHY6FZZ, DHY77ZZ, DHY7FZZ, DHY87ZZ, DHY8FZZ, DHY97ZZ, DHY9FZZ, DHYB7ZZ, DHYBFZZ, DHYCFZZ, DM000ZZ, DM001ZZ, DM002ZZ, DM003Z0, DM003ZZ, DM004ZZ, DM005ZZ, DM010ZZ, DM011ZZ, DM012ZZ, DM013Z0, DM013ZZ, DM014ZZ, DM015ZZ, DM1097Z, DM1098Z, DM1099Z, DM109BZ, DM109CZ, DM109YZ, DM10B7Z, DM10B8Z, DM10B9Z, DM10BB1, DM10BBZ, DM10BCZ, DM10BYZ, DM1197Z, DM1198Z, DM1199Z, DM119BZ, DM119CZ, DM119YZ, DM11B7Z, DM11B8Z, DM11B9Z, DM11BB1, DM11BBZ, DM11BCZ, DM11BYZ, DMY07ZZ, DMY0FZZ, DMY17ZZ, DMY1FZZ, DP000ZZ, DP001ZZ, DP002ZZ, DP003Z0, DP003ZZ, DP004ZZ, DP005ZZ, DP020ZZ, DP021ZZ, DP022ZZ, DP023Z0, DP023ZZ, DP024ZZ, DP025ZZ, DP030ZZ, DP031ZZ, DP032ZZ, DP033Z0, DP033ZZ, DP034ZZ, DP035ZZ, DP040ZZ, DP041ZZ, DP042ZZ, DP043Z0, DP043ZZ, DP044ZZ, DP045ZZ, DP050ZZ, DP051ZZ, DP052ZZ, DP053Z0, DP053ZZ, DP054ZZ, DP055ZZ, DP060ZZ, DP061ZZ, DP062ZZ, DP063Z0, DP063ZZ, DP064ZZ, DP065ZZ, DP070ZZ, DP071ZZ, DP072ZZ, DP073Z0, DP073ZZ, DP074ZZ, DP075ZZ, DP080ZZ, DP081ZZ, DP082ZZ, DP083Z0, DP083ZZ, DP084ZZ, DP085ZZ, DP090ZZ, DP091ZZ, DP092ZZ, DP093Z0, DP093ZZ, DP094ZZ, DP095ZZ, DP0B0ZZ, DP0B1ZZ, DP0B2ZZ, DP0B3Z0, DP0B3ZZ, DP0B4ZZ, DP0B5ZZ, DP0C0ZZ, DP0C1ZZ, DP0C2ZZ, DP0C3Z0, DP0C3ZZ, DP0C4ZZ, DP0C5ZZ, DPY07ZZ, DPY0FZZ, DPY27ZZ, DPY2FZZ, DPY37ZZ, DPY3FZZ, DPY47ZZ, DPY4FZZ, DPY57ZZ, DPY5FZZ, DPY67ZZ, DPY6FZZ, DPY77ZZ, DPY7FZZ, DPY87ZZ, DPY8FZZ, DPY97ZZ, DPY9FZZ, DPYB7ZZ, DPYBFZZ, DPYC7ZZ, DPYCFZZ, DT000ZZ, DT001ZZ, DT002ZZ, DT003Z0, DT003ZZ, DT004ZZ, DT005ZZ, DT010ZZ, DT011ZZ, DT012ZZ, DT013Z0, DT013ZZ, DT014ZZ, DT015ZZ, DT020ZZ, DT021ZZ, DT022ZZ, DT023Z0, DT023ZZ, DT024ZZ, DT025ZZ, DT030ZZ, DT031ZZ, DT032ZZ, DT033Z0, DT033ZZ, DT034ZZ, DT035ZZ, DT1097Z, DT1098Z, DT1099Z, DT109BZ, DT109CZ, DT109YZ, DT10B7Z, DT10B8Z, DT10B9Z, DT10BB1, DT10BBZ, DT10BCZ, DT10BYZ, DT1197Z, DT1198Z, DT1199Z, DT119BZ, DT119CZ, DT119YZ, DT11B7Z, DT11B8Z, DT11B9Z, DT11BB1, DT11BBZ, DT11BCZ, DT11BYZ, DT1297Z, DT1298Z, DT1299Z, DT129BZ, DT129CZ, DT129YZ, DT12B7Z, DT12B8Z, DT12B9Z, DT12BB1, DT12BBZ, DT12BCZ, DT12BYZ, DT1397Z, DT1398Z, DT1399Z, DT139BZ, DT139CZ, DT139YZ, DT13B7Z, DT13B8Z, DT13B9Z, DT13BB1, DT13BBZ, DT13BCZ, DT13BYZ, DTY07ZZ, DTY0CZZ, DTY0FZZ, DTY17ZZ, DTY1CZZ, DTY1FZZ, DTY27ZZ, DTY2CZZ, DTY2FZZ, DTY37ZZ, DTY3CZZ, DTY3FZZ, DU000ZZ, DU001ZZ, DU002ZZ, DU003Z0, DU003ZZ, DU004ZZ, DU005ZZ, DU010ZZ, DU011ZZ, DU012ZZ, DU013Z0, DU013ZZ, DU014ZZ, DU015ZZ, DU020ZZ, DU021ZZ, DU022ZZ, DU023Z0, DU023ZZ, DU024ZZ, DU025ZZ, DU1097Z, DU1098Z, DU1099Z, DU109BZ, DU109CZ, DU109YZ, DU10B7Z, DU10B8Z, DU10B9Z, DU10BB1, DU10BBZ, DU10BCZ, DU10BYZ, DU1197Z, DU1198Z, DU1199Z, DU119BZ, DU119CZ, DU119YZ, DU11B7Z, DU11B8Z, DU11B9Z, DU11BB1, DU11BBZ, DU11BCZ, DU11BYZ, DU1297Z, DU1298Z, DU1299Z, DU129BZ, DU129CZ, DU129YZ, DU12B7Z, DU12B8Z, DU12B9Z, DU12BB1, DU12BBZ, DU12BCZ, DU12BYZ, DUY07ZZ, DUY0CZZ, DUY0FZZ, DUY17ZZ, DUY1CZZ, DUY1FZZ, DUY27ZZ, DUY2CZZ, DUY2FZZ, DV000ZZ, DV001ZZ, DV002ZZ, DV003Z0, DV003ZZ, DV004ZZ, DV005ZZ, DV010ZZ, |

| | |
|---|---|
| | DV011ZZ, DV012ZZ, DV013Z0, DV013ZZ, DV014ZZ, DV015ZZ, DV1097Z, DV1098Z, DV1099Z, DV109BZ, DV109CZ, DV109YZ, DV10B7Z, DV10B8Z, DV10B9Z, DV10BB1, DV10BBZ, DV10BCZ, DV10BYZ, DV1197Z, DV1198Z, DV1199Z, DV119BZ, DV119CZ, DV119YZ, DV11B7Z, DV11B8Z, DV11B9Z, DV11BB1, DV11BBZ, DV11BCZ, DV11BYZ, DVY07ZZ, DVY0CZZ, DVY0FZZ, DVY17ZZ, DVY1FZZ, DW010ZZ, DW011ZZ, DW012ZZ, DW013Z0, DW013ZZ, DW014ZZ, DW015ZZ, DW020ZZ, DW021ZZ, DW022ZZ, DW023Z0, DW023ZZ, DW024ZZ, DW025ZZ, DW030ZZ, DW031ZZ, DW032ZZ, DW033Z0, DW033ZZ, DW034ZZ, DW035ZZ, DW040ZZ, DW041ZZ, DW042ZZ, DW043Z0, DW043ZZ, DW044ZZ, DW045ZZ, DW050ZZ, DW051ZZ, DW052ZZ, DW053Z0, DW053ZZ, DW054ZZ, DW055ZZ, DW060ZZ, DW061ZZ, DW062ZZ, DW063Z0, DW063ZZ, DW064ZZ, DW065ZZ, DW10BB1, DW10BBZ, DW1197Z, DW1198Z, DW1199Z, DW119BZ, DW119CZ, DW119YZ, DW11B7Z, DW11B8Z, DW11B9Z, DW11BB1, DW11BBZ, DW11BCZ, DW11BYZ, DW1297Z, DW1298Z, DW1299Z, DW129BZ, DW129CZ, DW129YZ, DW12B7Z, DW12B8Z, DW12B9Z, DW12BB1, DW12BBZ, DW12BCZ, DW12BYZ, DW1397Z, DW1398Z, DW1399Z, DW139BZ, DW139CZ, DW139YZ, DW13B7Z, DW13B8Z, DW13B9Z, DW13BB1, DW13BBZ, DW13BCZ, DW13BYZ, DW1697Z, DW1698Z, DW1699Z, DW169BZ, DW169CZ, DW169YZ, DW16B7Z, DW16B8Z, DW16B9Z, DW16BB1, DW16BBZ, DW16BCZ, DW16BYZ, DW1KBB1, DW1KBBZ, DW1LBB1, DW1LBBZ, DW1PBB1, DW1PBBZ, DW1QBB1, DW1QBBZ, DW1RBB1, DW1RBBZ, DW1XBB1, DW1XBBZ, DW1YBB1, DW1YBBZ, DWY17ZZ, DWY1FZZ, DWY27ZZ, DWY2FZZ, DWY37ZZ, DWY3FZZ, DWY47ZZ, DWY4FZZ, DWY57ZZ, DWY5FZZ, DWY5GDZ, DWY5GFZ, DWY5GGZ, DWY5GHZ, DWY5GYZ, DWY67ZZ, DWY6FZZ |

**Table S4. Classification of Surgery Type by Risk of Complications.**

| Surgical Risk Category | Codes |
|---|---|
| **Low** | **CPT:** 0581T, 28525, 44204, 47100, 47370, 50543, 50546, 50548<br><br>**ICD-9-CM:** 1732, 1733, 1735, 1736, 1742, 4382, 4522, 4581, 5025, 6553<br><br>**ICD-10-CM:** 06L28CZ, 06L28DZ, 06L28ZZ, 0B538ZZ, 0B548ZZ, 0B558ZZ, 0B568ZZ, 0B578ZZ, 0B588ZZ, 0B598ZZ, 0B5B8ZZ, 0B5K3ZZ, 0B5K4ZZ, 0B5K8ZZ, 0B5L3ZZ, 0B5L4ZZ, 0B5L8ZZ, 0B5M3ZZ, 0B5M4ZZ, 0B5M8ZZ, 0BQK3ZZ, 0BQK4ZZ, 0BQK8ZZ, 0BQL3ZZ, 0BQL4ZZ, 0BQL8ZZ, 0BQM3ZZ, 0BQM4ZZ, 0BQM8ZZ, 0C5R3ZZ, 0C5R4ZZ, 0C5R8ZZ, 0D15474, 0D15476, 0D15479, 0D1547A, 0D1547B, 0D154J4, 0D154J6, 0D154J9, 0D154JA, 0D154JB, 0D154K4, 0D154K6, 0D154K9, 0D154KA, 0D154KB, 0D154Z4, 0D154Z6, 0D154Z9, 0D154ZA, 0D154ZB, 0D15876, 0D15879, 0D1587A, 0D1587B, 0D158J6, 0D158J9, 0D158JA, 0D158JB, 0D158K6, 0D158K9, 0D158KA, 0D158KB, 0D158Z6, 0D158Z9, 0D158ZA, 0D158ZB, 0D16879, 0D1687A, 0D168J9, 0D168JA, 0D168K9, 0D168KA, 0D168Z9, 0D168ZA, 0D188KM, 0D188KN, 0D188KP, 0D188KQ, 0D188Z4, 0D188Z8, 0D188ZH, 0D188ZK, 0D188ZL, 0D188ZM, 0D188ZN, 0D188ZP, 0D188ZQ, 0D1B4ZQ, 0D1B8ZQ, 0D1E474, 0D1E47E, 0D1E47P, 0D1E4J4, 0D1E4JE, 0D1E4JP, 0D1E4K4, 0D1E4KE, 0D1E4KP, 0D1E4Z4, 0D1E4ZE, 0D1E4ZP, 0D1E874, 0D1E87E, 0D1E87P, 0D1E8J4, 0D1E8JE, 0D1E8JP, 0D1E8K4, 0D1E8KE, 0D1E8KP, 0D1E8Z4, 0D1E8ZE, 0D1E8ZP, 0DH83UZ, 0DH84UZ, 0DH87UZ, 0DH88UZ, 0DH93UZ, 0DH94UZ, 0DH97UZ, 0DH98UZ, 0DHA7UZ, 0DJD8ZZ, 0DX64Z5, 0DX84Z5, 0DXE4Z5, 0DXE4Z7, 0F503ZF, 0F503ZZ, 0F504ZF, 0F504ZZ, 0F513ZF, 0F514ZF, 0F523ZF, 0F524ZF, 0F5G3ZF, 0F5G4ZF, 0FQ03ZZ, 0FQ04ZZ, 0H5T3ZZ, 0H5T8ZZ, 0H5U3ZZ, 0H5U8ZZ, 0H5V3ZZ, 0H5V8ZZ, 0HHT3NZ, 0HHT7NZ, 0HHT8NZ, 0HHU3NZ, 0HHU7NZ, 0HHU8NZ, 0HHV3NZ, 0HHV7NZ, 0HHV8NZ, 0HHW3NZ, 0HHW7NZ, 0HHW8NZ, 0HHX3NZ, 0HHX7NZ, 0HHX8NZ, 0NQT3ZZ, 0NQT4ZZ, 0NQV3ZZ, 0NQV4ZZ, 0NRC47Z, 0NRC4KZ, 0NRD47Z, 0NRD4KZ, 0NRF47Z, 0NRF4KZ, 0NRG47Z, 0NRG4KZ, 0NRH47Z, 0NRH4KZ, 0NRJ47Z, 0NRJ4KZ, 0NRK47Z, 0NRK4KZ, 0NRL47Z, 0NRL4KZ, 0NRM47Z, 0NRM4KZ, 0NRN47Z, 0NRN4KZ, 0NUC47Z, 0NUC4KZ, 0NUD47Z, 0NUD4KZ, 0NUF47Z, 0NUF4KZ, 0NUG47Z, 0NUG4KZ, 0NUH47Z, 0NUH4KZ, 0NUJ47Z, 0NUJ4KZ, 0NUK47Z, 0NUK4KZ, 0NUL47Z, 0NUL4KZ, 0NUM47Z, 0NUM4KZ, 0NUN47Z, 0NUN4KZ, 0NUP47Z, 0NUP4KZ, 0NUQ47Z, 0NUQ4KZ, 0NUT47Z, 0NUT4JZ, 0NUT4KZ, 0NUV47Z, 0NUV4JZ, 0NUV4KZ, 0TUB47Z, 0TUB4JZ, 0TUB4KZ, 0TUB87Z, 0TUB8JZ, 0TUB8KZ, 0U508ZZ, 0U518ZZ, 0U528ZZ, 0U548ZZ, 0V508ZZ, 0WC43ZZ, 0WC44ZZ, 0WC53ZZ, 0WC54ZZ, 8E0W4CZ, XV508A4 |
| **Moderate** | **CPT:** 0546T, 11970, 19125, 19126, 19294, 19301, 19302, 19303, 19304, 19305, 19306, 19307, 19340, 19342, 19350, 19357, 19361, 19364, 19366, 19367, 19369, 19380, 21299, 21554, 21558, 21601, 21602, 21603, 31599, 42844, 42950, 43122, 43276, 43277, 44207, 45123, 47382, 48999, 50234, 50236, 50240, 52601, 52648, 53854, 55845, 55874, 58550, 58552, 58950, 58953<br><br>**ICD-9-CM:** 1739, 215, 2169, 252, 253, 254, 2742, 2757, 2933, 301, 3021, 3022, 3029, 303, 304, 3220, 3221, 3222, 3223, 3224, 3225, 3226, 3227, 3228, 3229, 3241, 3249, 4040, 4042, 4240, 4241, 4242, 4251, 4252, 4253, 4254, 4255, 4256, 4258, 4259, 4261, 4262, 4263, 4264, 4265, 4269, 4342, 4381, 4389, 4391, 4399, 4439, 4562, 4572, 4573, 4575, 4576, 4579, 4582, 4583, 4595, 4604, 4639, 4651, 4652, 4842, 4849, 496, 5023, 5024, 5026, 5029, 503, 5099, 527, 555, 5551, 5552, 5779, 5787, 6021, 6029, 603, 604, 605, 6069, 6551, 6552, 6561, 6562, 6831, 6839, 6841, 6849, 6851, 6861, 6869, 6871, 6879, 689, 7631, 7643, 7645, 7691, 8520, 8521, 8523, 8541, 8542, 8543, 8544, 8545, 8546, 8547, 8548, 8553, 8554, 8570, 8579, 8583, 8584, 8593, 8595, 8596<br><br>**ICD-10-CM:** 06L27CZ, 06L27DZ, 06L27ZZ, 09BL0ZZ, 09BL3ZZ, 09BL4ZZ, 09BL7ZZ, 09BL8ZZ, 09BM0ZZ, 09BM3ZZ, 09BM4ZZ, 09BN0ZZ, 09BN3ZZ, 09BN4ZZ, 09BN7ZZ, 09BN8ZZ, 0B5K0ZZ, 0B5K7ZZ, 0B5L0ZZ, 0B5L7ZZ, 0B5M0ZZ, 0B5M7ZZ, 0BBC4ZZ, 0BBD4ZZ, 0BBF4ZZ, 0BBG4ZZ, 0BBH4ZZ, 0BBJ4ZZ, 0BBK0ZZ, 0BBK3ZZ, 0BBK4ZZ, 0BBK7ZZ, 0BBK8ZZ, 0BBL0ZZ, 0BBL3ZZ, 0BBL4ZZ, 0BBL7ZZ, 0BBL8ZZ, 0BBM0ZZ, 0BBM3ZZ, 0BBM4ZZ, 0BBM7ZZ, 0BBM8ZZ, 0BQK0ZZ, 0BQK7ZZ, 0BQL0ZZ, 0BQL7ZZ, |

| | |
|---|---|
| | 0BQM0ZZ, 0BQM7ZZ, 0C5R0ZZ, 0C5R7ZZ, 0CB00ZZ, 0CB03ZZ, 0CB0XZZ, 0CB10ZZ, 0CB13ZZ, 0CB1XZZ, 0CB70ZZ, 0CB73ZZ, 0CB7XZZ, 0CBM0ZZ, 0CBM3ZZ, 0CBM4ZZ, 0CBM7ZZ, 0CBM8ZZ, 0CBR0ZZ, 0CBR3ZZ, 0CBR4ZZ, 0CBR7ZZ, 0CBR8ZZ, 0CBS0ZZ, 0CBS3ZZ, 0CBS4ZZ, 0CBS7ZZ, 0CBS8ZZ, 0CBT0ZZ, 0CBT3ZZ, 0CBT4ZZ, 0CBT7ZZ, 0CBT8ZZ, 0CBV0ZZ, 0CBV3ZZ, 0CBV4ZZ, 0CBV7ZZ, 0CBV8ZZ, 0CX00ZZ, 0CX0XZZ, 0CX10ZZ, 0CX1XZZ, 0CX40ZZ, 0CX4XZZ, 0CX50ZZ, 0CX5XZZ, 0CX60ZZ, 0CX6XZZ, 0D15074, 0D15076, 0D15079, 0D1507A, 0D1507B, 0D150J4, 0D150J6, 0D150J9, 0D150JA, 0D150JB, 0D150K4, 0D150K6, 0D150K9, 0D150KA, 0D150KB, 0D150Z4, 0D150Z6, 0D150Z9, 0D150ZA, 0D150ZB, 0D153J4, 0D16079, 0D1607A, 0D160J9, 0D160JA, 0D160K9, 0D160KA, 0D160Z9, 0D160ZA, 0D1B0ZQ, 0D1E074, 0D1E07E, 0D1E07P, 0D1E0J4, 0D1E0JE, 0D1E0JP, 0D1E0K4, 0D1E0KE, 0D1E0KP, 0D1E0Z4, 0D1E0ZE, 0D1E0ZP, 0DB50ZZ, 0DB53ZZ, 0DB57ZZ, 0DB60ZZ, 0DB63ZZ, 0DB64Z3, 0DB64ZZ, 0DB67ZZ, 0DB68ZZ, 0DBE0ZZ, 0DBE3ZZ, 0DBE4ZZ, 0DBE7ZZ, 0DBE8ZZ, 0DBF0ZZ, 0DBG0ZZ, 0DBK0ZZ, 0DBL0ZZ, 0DBM0ZZ, 0DBN0ZZ, 0DBP0ZZ, 0DBP4ZZ, 0DBQ0ZZ, 0DBQ3ZZ, 0DBQ4ZZ, 0DBQ7ZZ, 0DBQ8ZZ, 0DBQXZZ, 0DBR0ZZ, 0DBR3ZZ, 0DBR4ZZ, 0DH80UZ, 0DH90UZ, 0DHA0UZ, 0DX60Z5, 0DX80Z5, 0DXE0Z5, 0DXE0Z7, 0F500ZF, 0F500ZZ, 0F510ZF, 0F520ZF, 0F5G0ZF, 0FB00ZZ, 0FB03ZZ, 0FB04ZZ, 0FBG0ZZ, 0FBG3ZZ, 0FBG4ZZ, 0FQ00ZZ, 0H0T0JZ, 0H0T0KZ, 0H0T3JZ, 0H0T3KZ, 0H0U0JZ, 0H0U0KZ, 0H0U3JZ, 0H0U3KZ, 0H0V07Z, 0H0V0JZ, 0H0V0KZ, 0H0V37Z, 0H0V3JZ, 0H0V3KZ, 0H5T0ZZ, 0H5T7ZZ, 0H5TXZZ, 0H5U0ZZ, 0H5U7ZZ, 0H5UXZZ, 0H5V0ZZ, 0H5V7ZZ, 0H5VXZZ, 0HBT0ZZ, 0HBT3ZZ, 0HBT7ZZ, 0HBT8ZZ, 0HBTXZZ, 0HBU0ZZ, 0HBU3ZZ, 0HBU7ZZ, 0HBU8ZZ, 0HBUXZZ, 0HBV0ZZ, 0HBV3ZZ, 0HBV7ZZ, 0HBV8ZZ, 0HBVXZZ, 0HDT0ZZ, 0HDU0ZZ, 0HDV0ZZ, 0HDY0ZZ, 0HHT0NZ, 0HHU0NZ, 0HHV0NZ, 0HHW0NZ, 0HHX0NZ, 0HRT07Z, 0HRT0JZ, 0HRT0KZ, 0HRT3JZ, 0HRT3KZ, 0HRTX7Z, 0HRTXKZ, 0HRU07Z, 0HRU0JZ, 0HRU0KZ, 0HRU3JZ, 0HRU3KZ, 0HRUX7Z, 0HRUXKZ, 0HRVX7Z, 0HRVXKZ, 0HUT0JZ, 0HUT0KZ, 0HUT3JZ, 0HUT3KZ, 0HUU0JZ, 0HUU0KZ, 0HUU3JZ, 0HUU3KZ, 0HUV0JZ, 0HUV3JZ, 0HWT0JZ, 0HWT3JZ, 0HWU0JZ, 0HWU3JZ, 0HX5XZZ, 0NBT0ZZ, 0NBT3ZZ, 0NBT4ZZ, 0NBV0ZZ, 0NBV3ZZ, 0NBV4ZZ, 0NQT0ZZ, 0NQTXZZ, 0NQV0ZZ, 0NQVXZZ, 0NRC07Z, 0NRC0KZ, 0NRC37Z, 0NRC3KZ, 0NRD07Z, 0NRD0KZ, 0NRD37Z, 0NRD3KZ, 0NRF07Z, 0NRF0KZ, 0NRF37Z, 0NRF3KZ, 0NRG07Z, 0NRG0KZ, 0NRG37Z, 0NRG3KZ, 0NRH07Z, 0NRH0KZ, 0NRH37Z, 0NRH3KZ, 0NRJ07Z, 0NRJ0KZ, 0NRJ37Z, 0NRJ3KZ, 0NRK07Z, 0NRK0KZ, 0NRK37Z, 0NRK3KZ, 0NRL07Z, 0NRL0KZ, 0NRL37Z, 0NRL3KZ, 0NRM07Z, 0NRM0KZ, 0NRM37Z, 0NRM3KZ, 0NRN07Z, 0NRN0KZ, 0NRN37Z, 0NRN3KZ, 0NRP0KZ, 0NRP3KZ, 0NRP4KZ, 0NRQ0KZ, 0NRQ3KZ, 0NRQ4KZ, 0NUC07Z, 0NUC0KZ, 0NUC37Z, 0NUC3KZ, 0NUD07Z, 0NUD0KZ, 0NUD37Z, 0NUD3KZ, 0NUF07Z, 0NUF0KZ, 0NUF37Z, 0NUF3KZ, 0NUG07Z, 0NUG0KZ, 0NUG37Z, 0NUG3KZ, 0NUH07Z, 0NUH0KZ, 0NUH37Z, 0NUH3KZ, 0NUJ07Z, 0NUJ0KZ, 0NUJ37Z, 0NUJ3KZ, 0NUK07Z, 0NUK0KZ, 0NUK37Z, 0NUK3KZ, 0NUL07Z, 0NUL0KZ, 0NUL37Z, 0NUL3KZ, 0NUM07Z, 0NUM0KZ, 0NUM37Z, 0NUM3KZ, 0NUN07Z, 0NUN0KZ, 0NUN37Z, 0NUN3KZ, 0NUP07Z, 0NUP0KZ, 0NUP37Z, 0NUP3KZ, 0NUQ07Z, 0NUQ0KZ, 0NUQ37Z, 0NUQ3KZ, 0NUT07Z, 0NUT0JZ, 0NUT0KZ, 0NUT37Z, 0NUT3JZ, 0NUT3KZ, 0NUV07Z, 0NUV0JZ, 0NUV0KZ, 0NUV37Z, 0NUV3JZ, 0NUV3KZ, 0TRB07Z, 0TRB0JZ, 0TRB0KZ, 0TRB47Z, 0TRB4JZ, 0TRB4KZ, 0TRB77Z, 0TRB7JZ, 0TRB7KZ, 0TRB87Z, 0TRB8JZ, 0TRB8KZ, 0TUB07Z, 0TUB0JZ, 0TUB0KZ, 0TUB77Z, 0TUB7JZ, 0TUB7KZ, 0V507ZZ, 0VB07ZZ, 0VB08ZZ, 0WC40ZZ, 0WC50ZZ, 0WQFXZ2 |
| **High** | **CPT:** 11971, 19120, 31420, 41120, 42890, 44120, 44140, 47120, 50220, 50225, 50230, 50545, 58548<br><br>**ICD-9-CM:** 1731, 1734, 214, 3230, 3239, 3250, 3259, 4571, 4574, 4850, 4851, 4852, 4859, 4862, 4863, 4865, 4869, 5022, 504, 5059, 5252, 5253, 526, 5771, 6549, 688, 8522, 8594<br><br>**ICD-10-CM:** 07T10ZZ, 07T14ZZ, 07T20ZZ, 07T24ZZ, 09TK0ZZ, 09TK4ZZ, 09TKXZZ, 09TL0ZZ, 09TL4ZZ, 09TL7ZZ, 09TL8ZZ, 09TM0ZZ, 09TM4ZZ, 09TN0ZZ, 09TN4ZZ, 09TN7ZZ, 09TN8ZZ, 0BTC0ZZ, 0BTC4ZZ, 0BTD0ZZ, 0BTD4ZZ, 0BTF0ZZ, 0BTF4ZZ, 0BTG0ZZ, 0BTG4ZZ, 0BTH4ZZ, 0BTJ0ZZ, 0BTJ4ZZ, 0BTK0ZZ, 0BTK4ZZ, 0BTL0ZZ, 0BTL4ZZ, 0BTM0ZZ, 0BTM4ZZ, 0CT70ZZ, 0CT7XZZ, 0CTM0ZZ, 0CTM4ZZ, 0CTM7ZZ, 0CTM8ZZ, 0CTR0ZZ, 0CTR4ZZ, 0CTR7ZZ, 0CTR8ZZ, 0CTS0ZZ, 0CTS4ZZ, 0CTS7ZZ, 0CTS8ZZ, 0CTT0ZZ, 0CTT4ZZ, 0CTT7ZZ, 0CTT8ZZ, 0CTV0ZZ, 0CTV4ZZ, 0CTV7ZZ, 0CTV8ZZ, 0DT50ZZ, 0DT54ZZ, 0DT57ZZ, 0DT58ZZ, 0DT60ZZ, 0DT64ZZ, 0DT67ZZ, 0DT68ZZ, 0DT90ZZ, 0DT94ZZ, 0DT97ZZ, 0DT98ZZ, 0DTA0ZZ, 0DTA4ZZ, 0DTA7ZZ, |

| | |
|---|---|
| | 0DTA8ZZ, 0DTB0ZZ, 0DTB4ZZ, 0DTB7ZZ, 0DTB8ZZ, 0DTE0ZZ, 0DTE4ZZ, 0DTE7ZZ, 0DTE8ZZ, 0DTF0ZZ, 0DTF4ZZ, 0DTF7ZZ, 0DTF8ZZ, 0DTG0ZZ, 0DTG4ZZ, 0DTG7ZZ, 0DTG8ZZ, 0DTGFZZ, 0DTH0ZZ, 0DTH4ZZ, 0DTH7ZZ, 0DTH8ZZ, 0DTK0ZZ, 0DTL0ZZ, 0DTL4ZZ, 0DTL7ZZ, 0DTL8ZZ, 0DTLFZZ, 0DTMFZZ, 0DTN0ZZ, 0DTN4ZZ, 0DTN7ZZ, 0DTN8ZZ, 0DTNFZZ, 0DTP0ZZ, 0DTP4ZZ, 0DTP7ZZ, 0DTP8ZZ, 0DTQ0ZZ, 0DTQ4ZZ, 0DTQ7ZZ, 0DTQ8ZZ, 0DTR0ZZ, 0DTR4ZZ, 0FT00ZZ, 0FT04ZZ, 0FT10ZZ, 0FT14ZZ, 0FT20ZZ, 0FT24ZZ, 0FTG0ZZ, 0FTG4ZZ, 0FY00Z0, 0FY00Z1, 0FY00Z2, 0HPT0JZ, 0HPT0NZ, 0HPT3JZ, 0HPT3NZ, 0HPU0JZ, 0HPU0NZ, 0HPU3JZ, 0HPU3NZ, 0HTT0ZZ, 0HTU0ZZ, 0HTV0ZZ, 0KTH0ZZ, 0KTJ0ZZ, 0NTC0ZZ, 0NTD0ZZ, 0NTF0ZZ, 0NTG0ZZ, 0NTH0ZZ, 0NTJ0ZZ, 0NTK0ZZ, 0NTL0ZZ, 0NTM0ZZ, 0NTN0ZZ, 0NTP0ZZ, 0NTQ0ZZ, 0NTR0ZZ, 0NTS0ZZ, 0RTC0ZZ, 0RTD0ZZ, 0TT00ZZ, 0TT04ZZ, 0TT10ZZ, 0TT14ZZ, 0TTB0ZZ, 0TTB4ZZ, 0TTB7ZZ, 0TTB8ZZ, 0TTD0ZZ, 0TTD4ZZ, 0TTD7ZZ, 0TTD8ZZ, 0UT00ZZ, 0UT07ZZ, 0UT08ZZ, 0UT0FZZ, 0UT10ZZ, 0UT17ZZ, 0UT18ZZ, 0UT1FZZ, 0UT20ZZ, 0UT24ZZ, 0UT27ZZ, 0UT28ZZ, 0UT2FZZ, 0UT40ZZ, 0UT44ZZ, 0UT70ZZ, 0UT90ZL, 0UT90ZZ, 0UT94ZL, 0UT94ZZ, 0UT97ZL, 0UT97ZZ, 0UT98ZL, 0UT98ZZ, 0UT9FZL, 0UT9FZZ, 0UTC0ZZ, 0UTC4ZZ, 0UTC7ZZ, 0UTC8ZZ, 0UTG0ZZ, 0VT00ZZ, 0VT04ZZ, 0VT07ZZ, 0VT08ZZ, 0VT30ZZ, 0VT34ZZ |

**Supplemental Figure 1. Number of surgeries by site of surgery for each cancer type among Medicaid beneficiaries with a first observed cancer surgery from 2001 – 2021.**

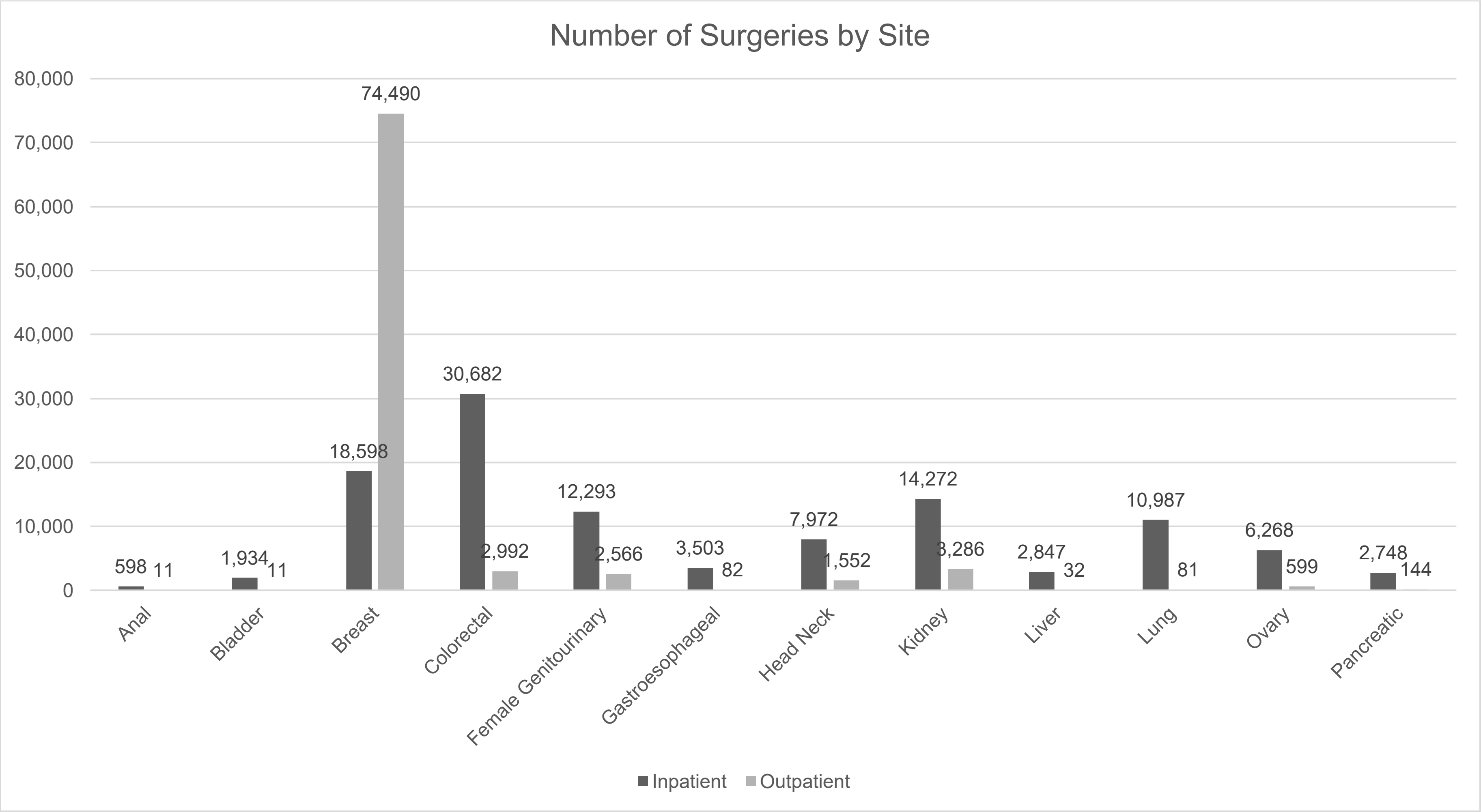
